# Acupuncture sets of trial reported core outcomes (ASTRO) for women’s health across the lifespan

**DOI:** 10.1101/2025.09.07.25335251

**Authors:** Kate M Levett, Valentina Buay, Claudia Citkovitz, Kathleen Lumiere, Rosa Schnyer, Sandro Graca, Debra Betts, Wen Tu, Belinda “Beau” Anderson, Lisa Conboy, Lisa Taylor-Swanson

## Abstract

**Introduction:** Clinical trials of acupuncture are difficult to meaningfully aggregate due to small sample sizes, and reductionist, heterogenous outcome measures. To improve the quality of evidence, standardized reporting of minimum data that reflects whole-person health, essential to East Asian Medicine (EAM), is needed. The Core Outcome Measures in Effectiveness Trials (COMET) initiative (1) offers a methodology for developing core outcome sets (COS) through collaborative group consensus, including consumers. This study presents the ‘Acupuncture Sets of Trial Reported Outcomes’ (ASTRO) for women’s health. Using domains of pain, sleep, mood/cognition, and safety, the ASTRO framework captures EAM complexity as well as standardizing reporting outcomes.

**Purpose:** To develop a COS for acupuncture trials in women’s health, reflecting whole-person health outcomes across the lifespan.

**Methods:** Following COMET guidelines, we used a three-stage consensus process involving practitioners, researchers, patients, and policymakers to develop a COS for reporting minimum data for acupuncture trials in women’s health. This process involved a systematic review, roundtable discussion, two-round Delphi survey, and a consensus meeting. We examined the conditions of menstrual health, PCOS, fertility, pregnancy, and menopause to develop an outcome set for safety, feasibility and EAM specific outcomes.

**Results:** From 320 studies, 579 outcomes were identified. A final COS of 12 outcomes was agreed upon:

- Safety (n=2): Serious adverse events (SAEs) and minor adverse events (AEs);
- EAM (n=8): Changes to primary and secondary patient reported outcome measures (PROMs), change in EAM diagnostic indices, patient experience/satisfaction, demographics, mood/cognition, sleep, and pain;
- Feasibility (n=2): Acceptability and cost.

Definitions for S/AEs follow Witt *et al.* (2) and Bäumler *et al.* (3), and outcomes are to be used in conjunction with those specified in the STRICTA guidelines (4).

**Conclusion:** The ASTRO Women’s Health COS enables consistent, whole-person outcome reporting, to improve the quality and comparability of acupuncture research.

**Summary box:** 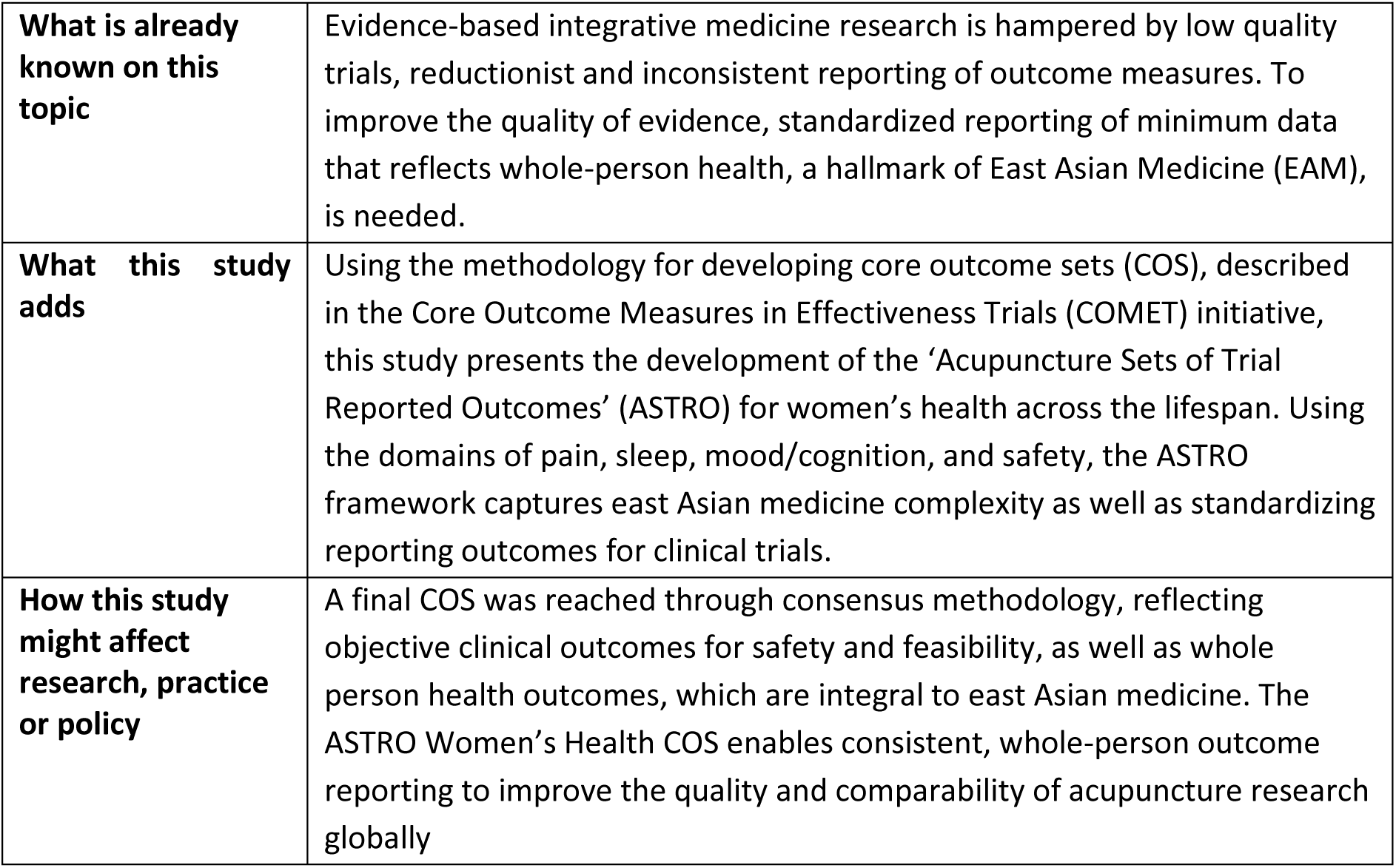

**Definitions of terms:** 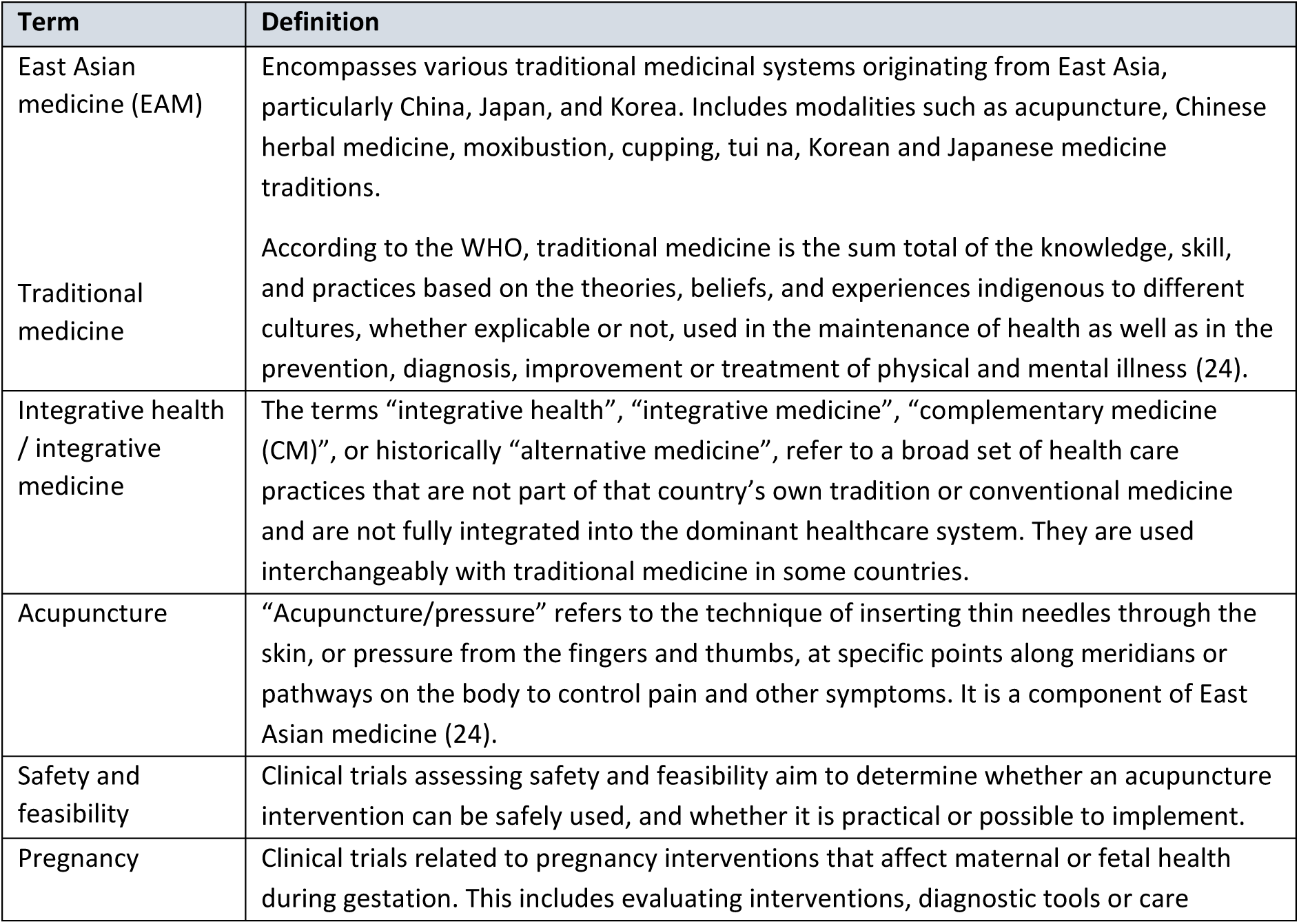

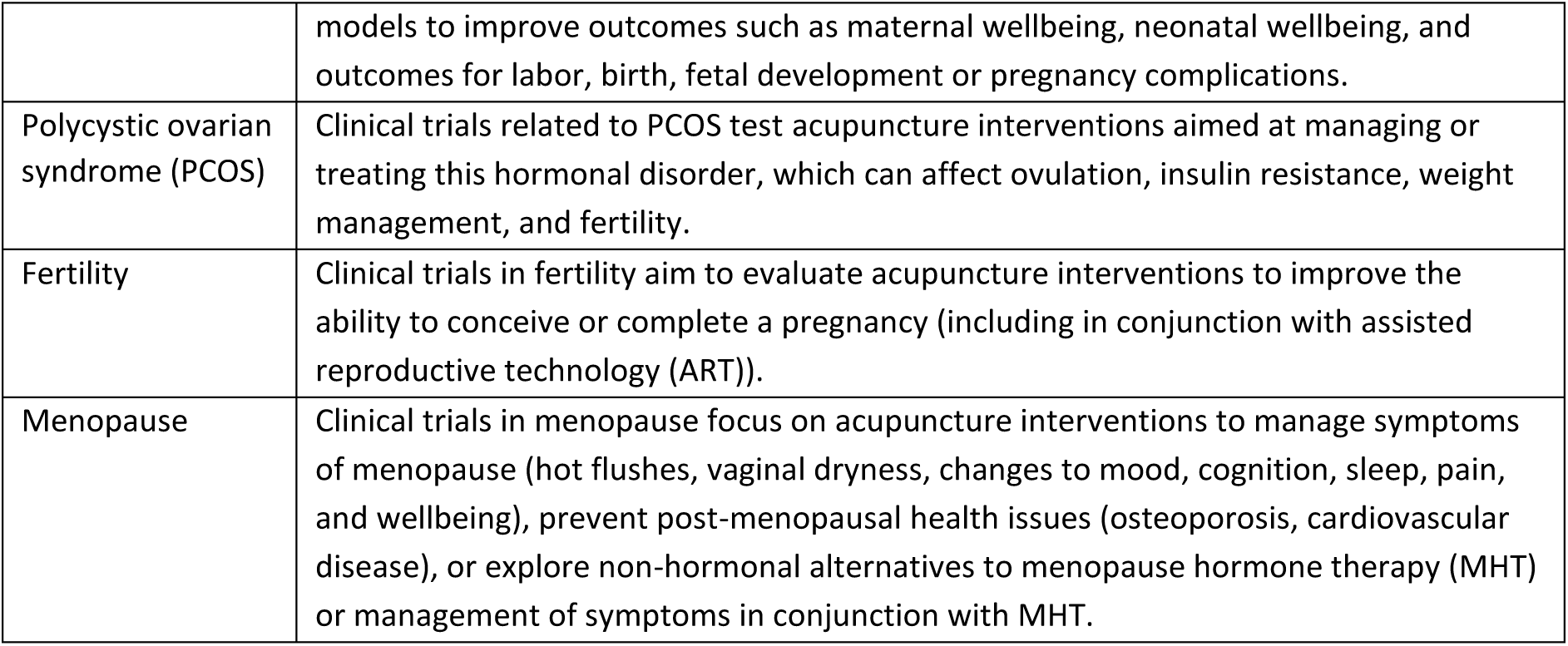

## Introduction

Research suggests that globally, women are the most frequent users of integrative therapies and traditional medicines, including acupuncture (5, 6). Although clinical trials and systematic reviews demonstrate the capacity for acupuncture to positively impact healthcare practice and integration, research in this area has been severely limited by issues such as methodological quality, low sample sizes and heterogenous outcome measures that have variable relevance to whole person health (8–12). Acupuncture, as part of traditional Chinese medicine (TCM) and other systems of east Asian medicine (EAM), holds whole person health as a core concept, emphasizing the important intersection of physical, mental, emotional, and environmental aspects of health. Women, particularly during key hormonal transitions such as menstruation, fertility, pregnancy, and menopause, are reported to be the most common users of acupuncture (13). However, researchers face significant challenges due to inconsistent reporting of outcome measures for acupuncture interventions. This heterogeneity makes it difficult to synthesize evidence and conduct meaningful meta-analyses to guide policy and practice, particularly in low resource settings (14–17). There is a strong need for standardized methods to assess the benefits and risks of acupuncture treatment for women across the lifespan, which would help inform practitioners, policymakers, and the public across the world (13–16).

Research indicates that acupuncture is effective for various aspects of women’s health conditions across the lifespan (18), including menstrual health disorders (19), PCOS (20), fertility (19), pregnancy-related conditions (21), and menopause (22). High-level evidence also supports the general safety of acupuncture (2, 23, 24). The mechanisms behind acupuncture’s effects are complex, and involve cross-cultural understanding of terminology and concepts, and ongoing debates about how best to measure outcomes (23). Foundational texts like the *Huang Di Nei Jing*, the oldest recorded acupuncture text dating back to the 4^th^-2^nd^ century BCE, have existed since antiquity, and yet continue to shape the philosophy and practice of EAM today (25, 26). These texts emphasize the integration of essential practices and whole person health philosophy in the use of EAM for healthcare. In more modern literature, acupuncture research has benefitted from advances in imaging and biological techniques, resulting in strong evidence for western medical biological and physiological pathways being validated for EAM. Acupuncture impacts multiple physiological systems including neuroendocrine regulation (27, 28), autonomic nervous system and immune system modulation (29, 30), pain modulation (31), and hormonal balancing (27, 32).

Current research also investigates how factors such as dosage, timing and methods of application influence the effectiveness of acupuncture treatments (27). Highlighting these factors as key research outcomes moves the research paradigm beyond the reductionist sham-controlled trial debate (33–35) and situates acupuncture research within the broader context of integrative whole person health (7). That said, measuring outcomes that truly reflect whole person health remains difficult and yet critically important in acupuncture research. Clinical practice of acupuncture in healthcare recognizes that for many women’s health conditions, patients present with symptom clusters. However, existing literature often reflects a reductionist approach, focusing only on a primary symptom or outcome. These outcomes are often too different between studies to draw meaningful conclusions in systematic reviews or meta-analyses and many fail to reflect patient-centered outcomes of interest. Additionally, although research generally supports the safety of acupuncture; however, systematic and condition-specific reporting of safety data is often lacking and does not always address the concerns of consumers, clinicians and policy makers (36).

Critical evaluation of clinical trials and systematic reviews should provide actionable evidence to improve integrated whole person health care for women (15). Creating COSs that reflect whole person health for women, through input from healthcare consumers, practitioners, and researchers, can help ensure that research outcomes are relevant to practice, policy, and consumer needs. This project will describe the concept for developing the ASTRO framework, for COS in acupuncture trials across the lifespan of women’s health, using a modified Delphi-technique for data harmonization and consensus gathering.

The aim of this study was to reach a robust consensus about a COS that reflects whole person health to be used when studying and evaluating interventions of acupuncture for women’s health and decision making for treatment options within and alongside conventional health services.

We employed a lifespan approach to propose common data elements for women’s health in the domains of *pain*, *sleep*, *mood*/*cognition*, and *safety*. This ASTRO framework reflects the complexity of women’s issues and emphasizes whole person health (7). We recommend using this ASTRO Women’s Health COS as a minimum standard to improve reporting across future studies of acupuncture studies, to reduce variation in outcome reporting and allow for better data synthesis.

### Research Question

In clinical studies of acupuncture for women’s health, can data harmonization and consensus building techniques produce a standardized COS of minimum reportable data that reflects the complexity and whole person health principles inherent to systems of EAM?

### Aims

This study aims to:

1. Use consensus methods to gather outcome data from the literature, roundtable discussions, online Delphi-style surveys and expert consensus meetings.
2. Identify core outcomes, to be used as the minimum standard for reporting in clinical studies of acupuncture for women’s health across the lifespan.
3. Develop a model for reporting core outcomes for acupuncture interventions for women’s health that reflects the complexity inherent in the principles of EAM.

### Inclusivity statement

The terms ‘women’, ‘maternal’, ‘mother’ or ‘obstetric’ are intended to include individuals who may not identify as women but who are, have been, or could have reproductive health concerns, become pregnant, or experience menopausal symptoms.

### Study scope and registration

This study was developed according to the COMET Handbook (35) and the protocol was registered with the COMET database (https://www.comet-initiative.org/). In accordance with the COMET handbook, our study involved four stages:

i) Systematic literature review to identify outcomes in existing literature
ii) Roundtable discussion of findings at a relevant international conference,
iii) Two-round modified Delphi survey with patients/consumers, clinicians, researchers, and other relevant stakeholders with an interest in women’s health,
iv) Final expert consensus meeting to refine the COS.

This report was developed in accordance with the Core Outcome Set–STAndards for Reporting: The COS-STAR Statement (37).

#### Study Registration

The COS development protocol and study has been registered in the COMET database: (https://www.comet-initiative.org/Studies/Details/3476).

#### Population

Identified outcomes are intended to be applicable to clinical trials of acupuncture for women’s health, involving patients treated for women’s health conditions, regardless of length of condition or healthcare setting.

#### Intervention

Interventions reporting AEM outcomes include trials of acupuncture, acupressure, tui na, and moxibustion for women’s health conditions including menstrual health conditions, fertility conditions, PCOS, pregnancy related conditions, and menopause.

### Ethics and consent

#### Ethics

Institutional ethics approval was obtained from the University of Notre Dame Australia’s Human Research Ethics Committee (024–078). Consent was obtained from all study participants by a click button entry to the survey. Survey data was anonymized, and any personal data was kept confidential. Only study investigators had access to the data via secure platforms.

#### Confidentiality

All participant responses were de-identified. Results are reported as aggregated responses with de-identified data where appropriate.

## Methods

This study reports an ASTRO Women’s Health COS using the methods described by the COMET initiative (38). We describe our methods for the development of the ASTRO framework which recommends minimum data to be reported for all acupuncture studies in women’s health, regardless of condition. We separately report the development of ASTRO extension COS for each of the five women’s health conditions (menstrual health, PCOS, fertility, pregnancy, and menopause). The ASTRO extensions for each condition are to be reported in addition to this primary ASTRO COS. Additionally, we searched the COMET database for existing studies of acupuncture and/or women’s health conditions, and our findings incorporate any outcomes identified in any previous COMET studies.

We used a multi-method design, as described by the COMET Initiative, involving a four-stage consensus building process including key collaborators: practitioners, researchers, patients, and policymakers, to develop a COS for reporting minimum data for acupuncture trials in women’s health. We engaged these key groups via conference proceedings at the Society for Acupuncture Research (SAR), professional organizations, and online groups, in the stages described in detail in the following section:

- **Stage I:** A systematic review of the literature across four databases identified outcome measures from acupuncture trials for five areas of women’s health (menstrual health conditions, fertility conditions, PCOS, pregnancy related conditions, and menopause).
- **Stage II:** Consultation with key collaborators via a roundtable discussion to refine measures for the development of survey items for the five women’s health surveys.
- **Stage III:** Surveys were distributed online to researcher, practice and consumer/patient networks, and conference attendees. Using a modified Delphi method, participants voted on each outcome’s importance (1 = not important, 9 = critically important). Outcomes averaging scores ≥7 were retained for a second-round survey, where the process was repeated.
- **Stage IV:** Items scoring ≥7 formed a final list, which were discussed at a final consensus meeting held at the SAR international research conference in 2025. Engaged stakeholders reached a final consensus to develop the COS for minimum reporting in trials of acupuncture for women’s health across the lifespan.

### Systematic literature search

Before conducting the systematic review, we searched the COMET trial database for studies in acupuncture in women’s health. Additionally, we registered our trial protocol to avoid duplication of efforts by other researchers. The COMET search strategy was drawn from recommendations within the COMET handbook (38) and by Gargon and colleagues (39). Where a study had been conducted in the specific health condition of interest, we noted the COS already recommended to determine similarities or differences for inclusion in our recommendations.

#### Information sources

We conducted a systematic literature review of MEDLINE (EBSCO), PubMed, CINAHL (EBSCO), and Cochrane Library (Wiley). The dates searched included all studies to date (April 2023). Search terms for search domains were employed via title and abstract only or title and abstract and key word. Limits were applied to studies in English, and human subjects. Details of the search strategy and study selection are provided in Appendix 1. This search was conducted until data saturation was reached, i.e., no new outcomes were identified.

We used the concepts “acupressure”, “acupuncture therapy”, “obstetric*”, “pain management” (methods), “analgesia”, “labo$r pain” (therapy) to inform search strategy using Medical Subject Headings (MeSH). MeSH check words included “female”, “humans”, “pregnan*” “pregnan#”, “reproduct*”, “gyn$ecology”. We note that the variations in usage of truncations and wildcards across databases (Appendix 1).

#### Organization of data

After identification of relevant texts and removal of duplicates, studies were screened, and outcomes were extracted and documented for each health condition using an excel spreadsheet. The data extracted included: i) study characteristics, ii) a comprehensive list of outcome measures used in each clinical trial, and iii) instruments used to measure core outcomes and methods for data collection. Outcomes were expressed in a matrix (Appendix 2).

From this complete outcome inventory, the group reached an agreement on the sub-categorization of outcomes into domains based on perceived relatedness (Table 4). From the team, two researchers worked on the list for each condition of interest to categorize the outcomes, with consensus from the group sought following completion of all items listed. Outcomes expressed that were similar in either wording or concept were grouped together. The lists were prepared for presentation and roundtable discussion at a post-conference workshop.

### Survey participants

Participants across the four stages of this study consisted of members of four different stakeholder groups:

i) Clinicians/practitioners providing care to women across their lifespan,
ii) Researchers interested in this field,
iii) Stakeholders/policymakers/health services involved in development and approval of women’s health policy,
iv) Patients/consumers with self-reported experience of the above conditions, or with an interest in EAM practices.

#### Recruitment

I. Participants in the roundtable discussion were attendees at the post-conference symposium of the Society for Acupuncture Research (SAR) International Research Conference on May 18 –21, 2023, in New York, United States of America (USA). Participants reviewed the data derived from the literature review and provided email addresses via a link to a Qualtrics (40) database if they wished to participate in the online survey rounds.
II. Participants who voted in the online modified-Delphi surveys rounds one and two were recruited from workshop attendees at the SAR conference 2023 in New York, and subsequently at the 2024 conference in Hong Kong, who provided email addresses via a link to a separate Qualtrics database. Participants were also recruited through online learning platforms and groups, professional networks, flyer distribution at conferences, social media groups, and acupuncture society membership lists (Table 2, Appendix 3). Participants were sent QR codes that, when scanned, provided links to the first and second Delphi surveys.
III. Participants in the final consensus meeting were attendees at a post-conference symposium workshop of the SAR International Research Conference on April 3-6, 2025 in Newport Beach, California, USA.

### Round table discussion workshop

The roundtable discussion was conducted at a post-conference workshop at the SAR International Research Conference in New York in May 2023. Using a ‘World Cafe’ method (41), this workshop brought together key stakeholders including clinicians, researchers, policymakers, and consumers in alignment with COMET guidelines (1, 38). Participants reviewed the findings of the systematic literature review in two rounds of discussion. In each round, participants discussed outcomes from the literature review in groups relating to one of five women’s health conditions (menstrual health, PCOS, fertility, pregnancy, and menopause). They participated in a maximum of two groups based on expertise and interest. During the workshop session, participants evaluated the relevance of the identified outcome measures, highlighted any significant outcomes that had not been captured, and determined which outcomes were clinically similar enough to be combined into a single measure. The insights and feedback gathered from this workshop were used to create the first round of Delphi surveys for online distribution.

### Delphi surveys

A Delphi method survey (42) was created in English, using Qualtrics® software (40) with the data gathered from the systematic review and roundtable discussion. It was piloted with a small group of researchers, and revised for accessibility, grammar, spelling, and clarity. The survey included an explanation of the study as well as plain English explanations of any medical or condition-specific terminology.

Outcomes were organized into subcategories, each presented on a separate page, with space that allowed respondents to rate multiple outcomes within that subcategory and suggest any additional outcomes they considered relevant. Respondents rated the importance of each outcome using a Likert scale (1 = not important, 5 = important but not critical, 9 = critically important). Demographic information, including stakeholder group, age category, gender identity, and country of residence, was also collected to inform the recruitment strategy and reporting.

#### Survey distribution

The first survey was open for responses from August to October 2024 and the second survey was open for responses from January to March 2025. We approached different groups where there was a connection to researchers’ interests, as well as online forums such as Reddit and Facebook (see below). Permission from group admins was sought where required. During this time, emails were sent for distribution, and reminder emails were sent at 48 and 24 hours before survey closure. We asked survey participants if they were willing to participate in the second survey round by leaving their email address via a link to a separate Qualtrics database so that responses remained anonymous.

##### Platforms and organizations used for recruitment of study participants

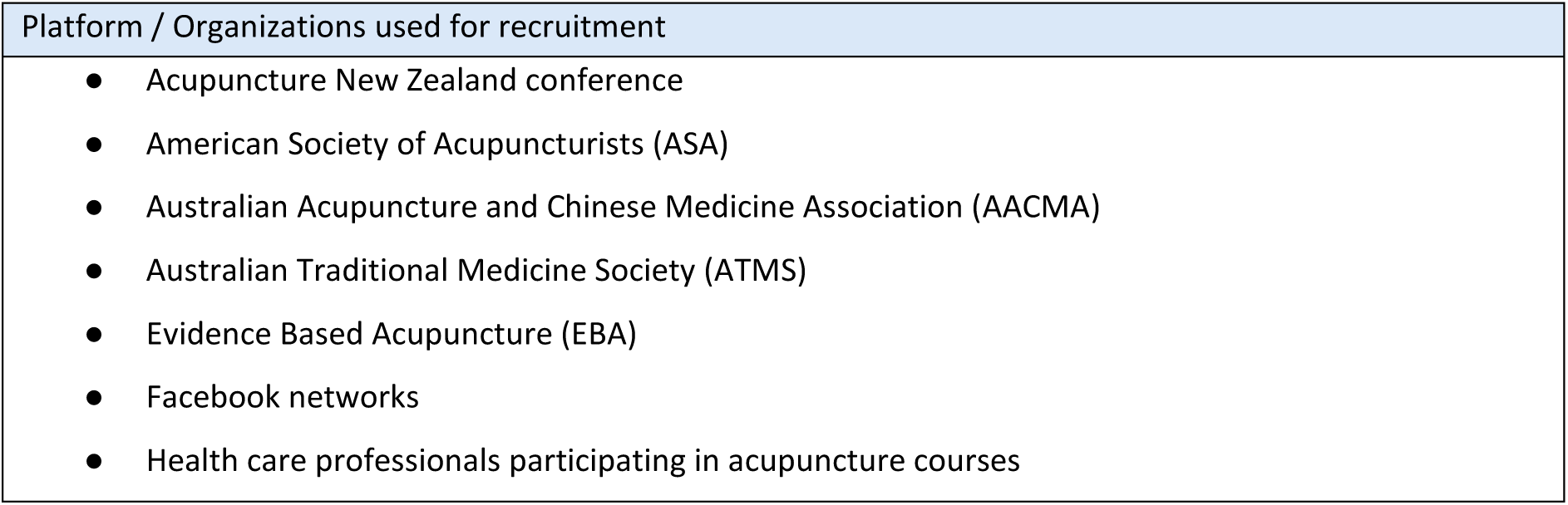
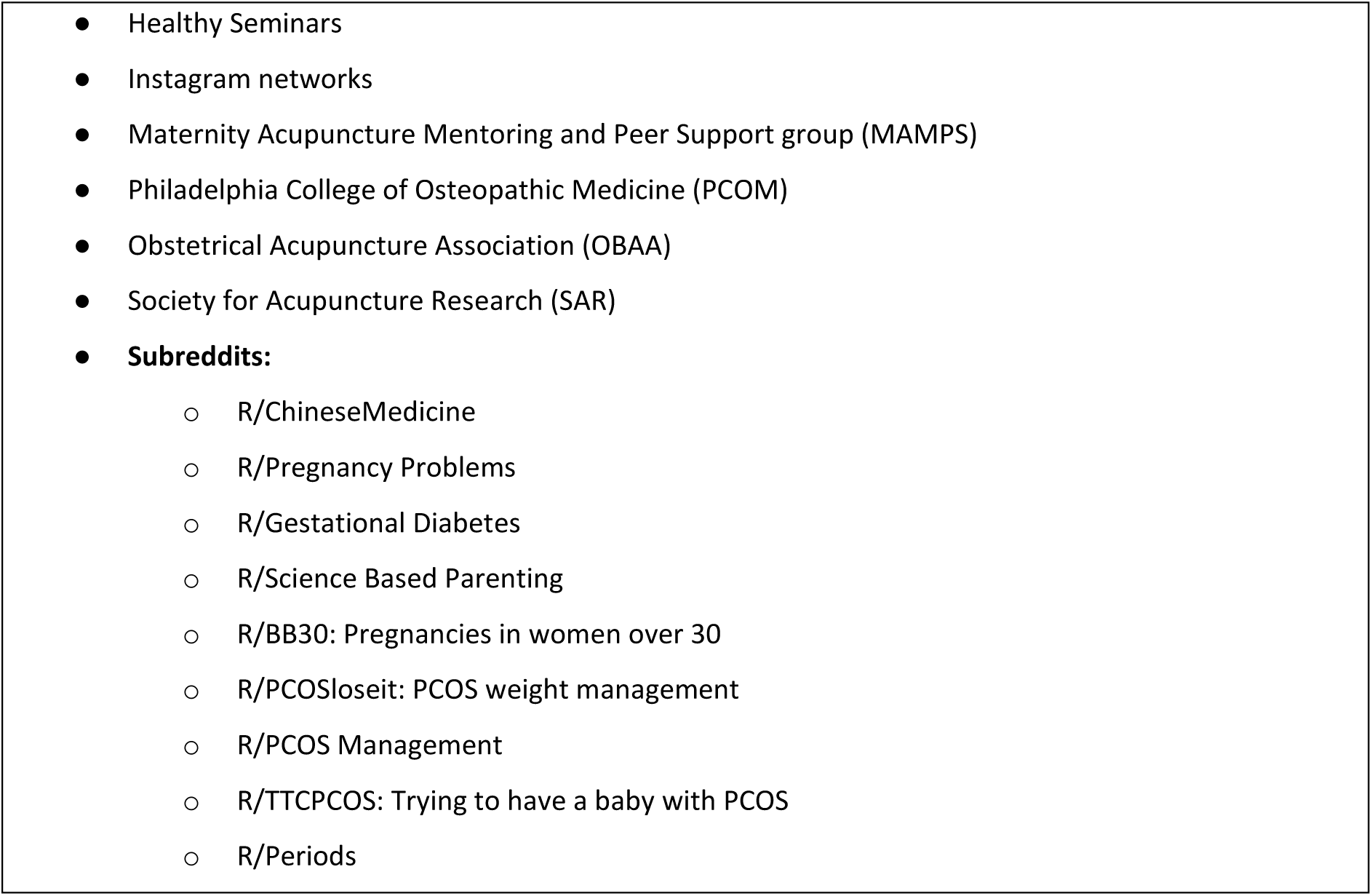

#### Survey data collection

Data from the completed first round surveys were downloaded from Qualtrics as a spreadsheet. Response scores for each outcome were averaged, and outcomes that scored an average importance rating of 7 or higher (out of 9) were retained for inclusion in the second survey. Respondents were given the opportunity to suggest outcomes they felt were important, including those previously removed or not yet considered. Scores from the first round were not provided to respondents in the subsequent round.

#### Data organization

To explore the safety, feasibility, and overall impact of acupuncture on women’s health, with a focus on whole-person health, the researchers organized the data into key domains: pain, sleep, mood/ cognition, EAM, and safety outcomes. This structure helped reduce complexity across each of the five survey sets.

After data collection from round two, survey responses were exported to a spreadsheet. Outcomes from all surveys which related to general EAM, safety, and feasibility of acupuncture were grouped into a primary common outcome set and were separated from those specific to each of the women’s health conditions. Two researchers for each survey grouped the outcomes by domain and average score, ranking them from highest to lowest. Based on these groupings, the team proposed six sets of core outcomes, one primary set, and five condition-specific sets. Each core set contained between six and 12 core outcomes for discussion at the consensus meeting. Outcomes that had been removed at earlier stages were flagged for possible re-inclusion. Additionally, outcomes identified in other COMET studies were presented to participants to provide context and to clarify why they were not included in the proposed core list, and to remain open for discussion.

### Consensus Meeting

The consensus meeting was conducted at a post conference workshop following the SAR 2025 conference, in California, USA. The workshop brought together 65 participants from over ten countries, representing a diverse group of acupuncture practitioners, researchers, policymakers, consumers, and advocates. A World Café approach (41) with two structured rounds was employed to facilitate collaborative discussion and consensus building. The process began with a plenary session presentation in which researchers reviewed the process of the COMET methodology and gave an update of progress to date following voting on outcomes for the two rounds of the surveys. Researchers introduced the proposed COS from the survey outcomes, and described the consensus methodology, meeting structure, and timeframes for the meeting. This was followed by an interactive question and answer period to ensure clarity and shared understanding among participants.

In the first round of discussions, the entire group discussed the EAM, safety, and feasibility outcomes, enabling broad input on these foundational outcome domains for the primary COS. For the second round, participants divided into smaller groups of two to 12 per group, each concentrating on one of five women’s health conditions (menstrual health, fertility, PCOS, pregnancy, or menopause), according to individual expertise and interest. Each group was facilitated by a researcher with condition-specific expertise, and participants were provided with printed documents listing the proposed outcomes (Appendix 3). Within these groups, participants annotated, discussed, and ranked outcomes, engaging in iterative dialogue until consensus was achieved for each condition-specific COS.

Through this process, participants debated and refined the outcome sets, ranking the outcomes and reaching consensus for the final COS (Table 6). Discussion ensured that the final selections reflected both the holistic principles of EAM and the requirements for biomedical measurability and clinical relevance. The high level of engagement and diverse perspectives contributed to the development of outcome measures that were meaningful and applicable to researchers, practitioners, and consumers alike. The results of the three stages of the COMET process to establish outcomes for minimally collected data will be presented below for the literature review, survey rounds and final consensus meeting by which the final COS was proposed.

## Results

### Initial list of outcomes

#### Literature review

Figure 1 illustrates the Preferred Reporting Items for Systematic Reviews and Meta-Analyses (PRISMA) flow chart for our systematic literature search. We searched databases (MEDLINE (EBSCO), PubMed, CINAHL (EBSCO), and Cochrane Library (Wiley)) for trials of acupuncture for each of the five women’s health conditions. The original search yielded 30,949 papers. After the removal of 19,004 duplicate papers, and a further 8672 irrelevant papers, 3,273 studies remained for screening. After abstract and title screening, 2,665 records were excluded due to wrong study design, wrong conditions, wrong intervention, or not being accessible in English. The remaining 608 were assessed for eligibility; with a further 282 studies deemed not eligible due to wrong modality, wrong study type, or inability for researchers to access full study. The search process shown in Figure 1, yielded a final 326 papers, reporting 579 separate outcomes (seen in Table 1).

**Figure 1.**
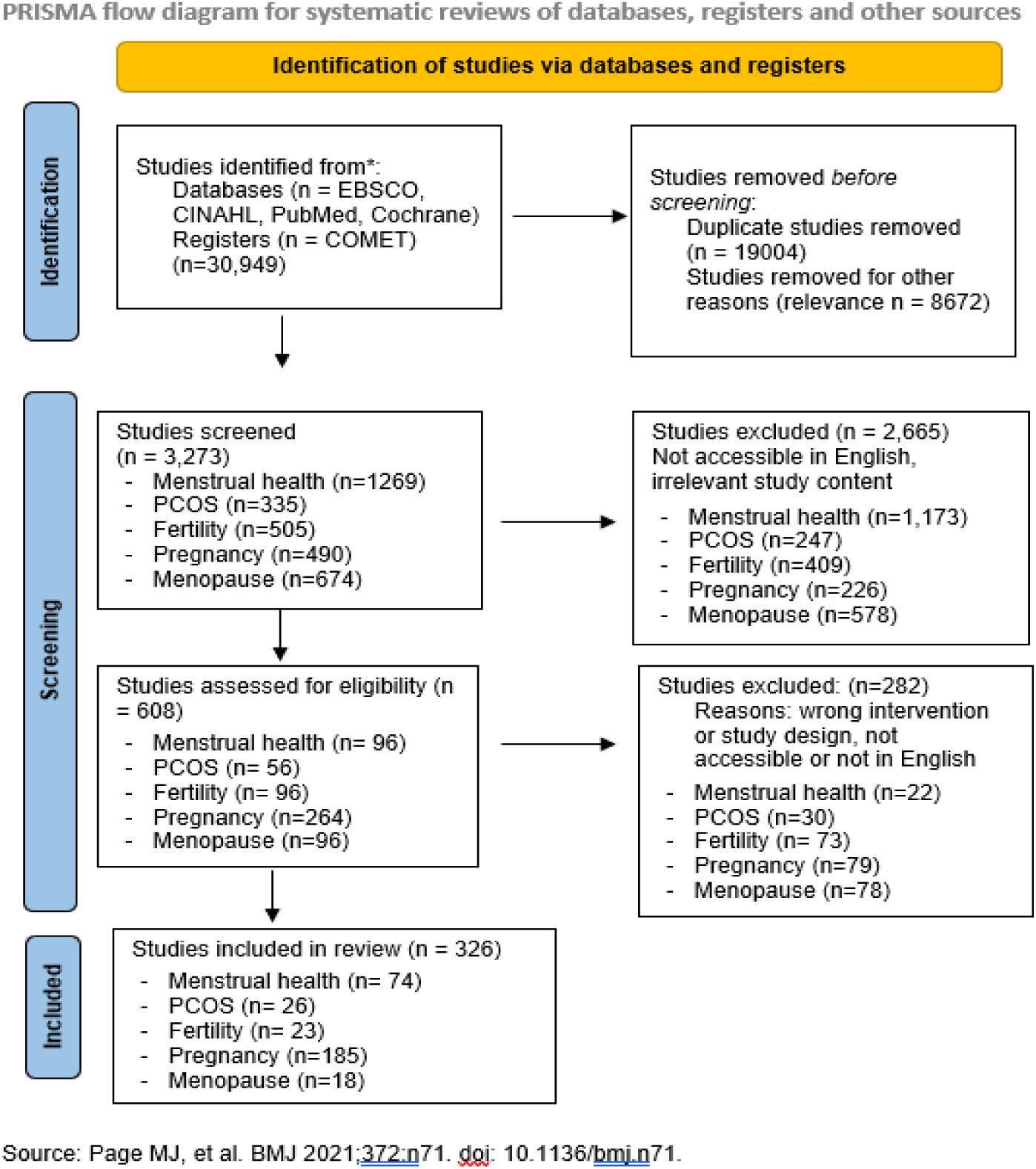
–Preferred Reporting Items for Systematic Reviews and Meta-Analyses (PRISMA) flow diagram of study selection.

##### COMET database search

We searched the COMET database to identify related studies and COS for acupuncture studies and/or women’s health condition specific studies. We identified two studies in pregnancy and childbirth which mention acupuncture as part of their treatment strategy. One Delphi study existed detailing expert consensus for breech version using moxibustion (43) and one Cochrane systematic review detailed pain relief methods in labour and birth (44). Both studies, while using different study designs, suggest using common outcomes for reporting in trials. The authors of the Cochrane review of pain in labour state that “Future trialists may, of course, wish to supplement these core outcome measures with additional topic-specific or trial-specific outcomes” (44).

Studies that followed the COMET methodology, with condition specific COS, but were not related to acupuncture or EAM, included one study for PCOS (45), three studies for menstrual health (46–48), one study for menopause (49), and two studies for general fertility (50, 51). For pregnancy studies, one study was for pelvic girdle pain (52), one for hyperemesis gravidarum (53), and one for multimorbidity in pregnancy (54). Where outcomes were already reported in published studies, we compared them to the ones arrived at in our study and presented the previous study outcomes at the consensus meeting for consideration.

##### Outcomes

From the final 326 papers that were assessed, 579 separate outcomes were listed across the five women’s health conditions as presented in Table 1. For a detailed list of outcomes under each domain please refer to Appendix 7. Researchers compiled the outcomes from all studies and combined those that had similar wording or meaning. The outcomes for pregnancy related conditions were the most numerous, reflecting the complexity and variability of outcomes in each pregnancy trimester, for labor and birth, and in the postpartum period. Pregnancy related outcomes were also captured in the fertility and PCOS studies.

**Table 1.** Papers and outcomes identified from literature search.

|  | Women's health condition | Papers (n) | Outcomes (n) |
| --- | --- | --- | --- |
| 1 | Menstrual health | 74 | 117 |
| 2 | PCOS | 26 | 94 |
| 3 | Fertility | 23 | 80 |
| 4 | Pregnancy | 185 | 171 |
| 5 | Menopause | 18 | 117 |
|  | **TOTAL** | **326** | **579** |

### Delphi Survey

Following the compilation of outcomes from the literature search, the first group of surveys were developed listing all outcomes, organized into domains and women’s health condition.

#### Organization of outcomes

Outcomes identified from the structured literature search for inclusion in round one of the Delphi surveys were further sub-categorized into 52 sub-domains across the five women’s health sets. Eight subcategories for acupuncture specific outcomes were identified which were relevant to all of the women’s health sets. These are;

| Outcome sub-categories for acupuncture outcomes |
| --- |
| ● Patient experience<br>● Pain<br>● Sleep<br>● Mood / Cognition<br>● Safety and adverse events<br>● Acupuncture/EAM specific outcomes<br>● Health condition severity and history<br>● Other |

Sub-categories for each of the individual surveys are shown in Appendix 6.

#### Participants

There were 319 participants in total for the two Delphi surveys, consisting of practitioners, researchers, policy makers and consumers/patients from over 20 countries around the world. Participants included conference attendees, online groups and participants from acupuncture societies and organizations. The majority of the 319 respondents were women (n=245, 77%), and identified as practitioners (n=192, 60%). Of the respondents, 34% came from the USA (n=107), 13% from Australia (n=40) and 12% from the United Kingdom (UK) (n=38). Table 2 shows the characteristics of the survey participants from surveys one and two. Appendix 4 and Appendix 5 further detail the respondent characteristics across each survey. In the first survey, 110 respondents identified as practitioners, 84 as patients/consumers, 13 as researchers, and nine as stakeholders/policymakers; for the second survey, 82, 21, 39, and 15 respondents identified as practitioners, patients, researchers, and policymakers respectively.

**Table 2.**
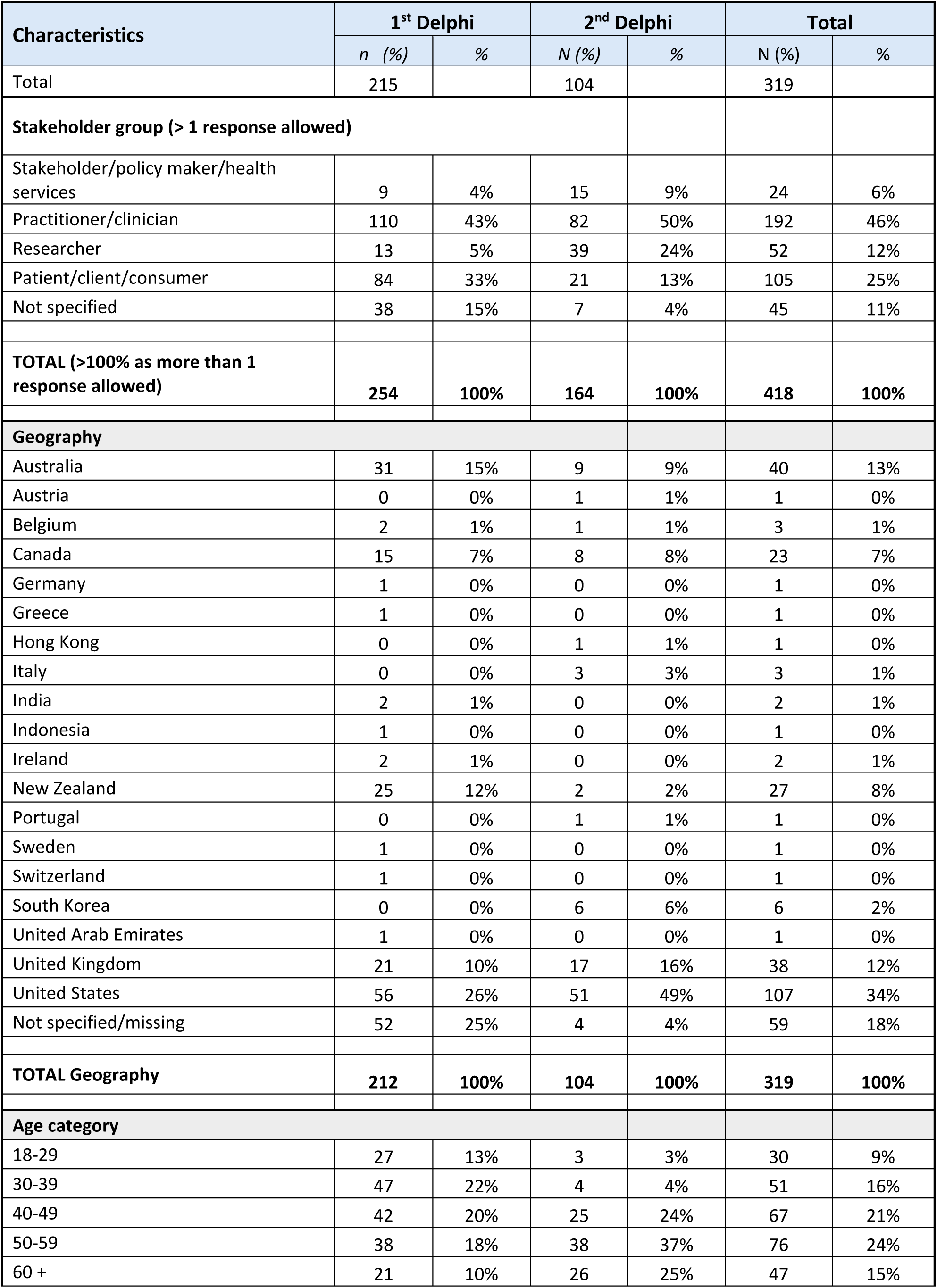
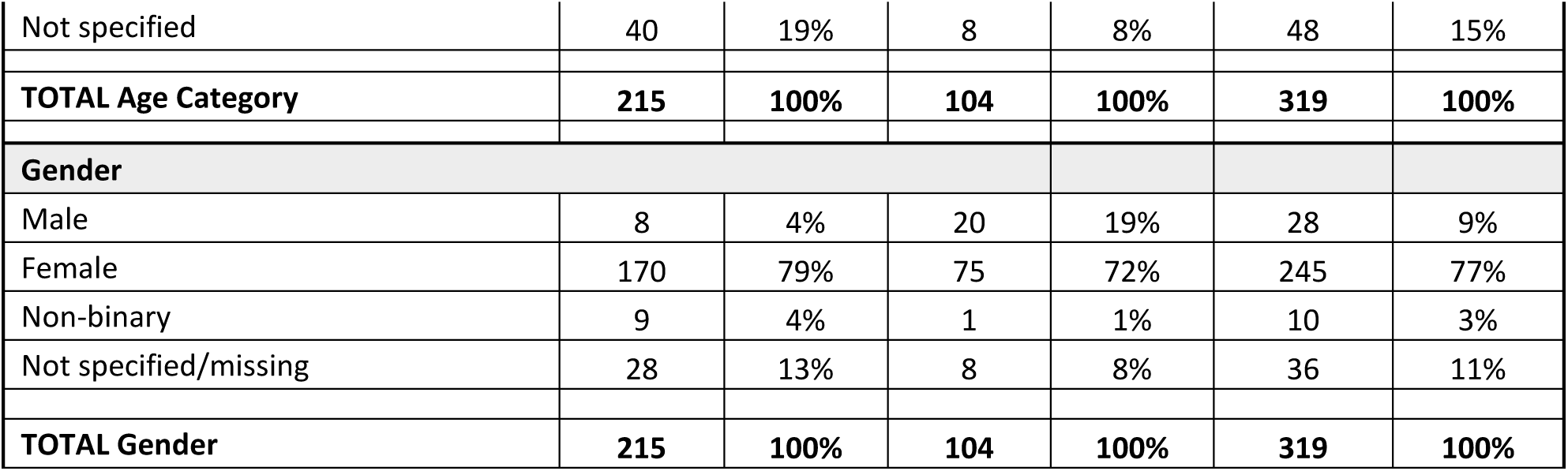
Characteristics of Delphi survey participants across all domains.

| Characteristics | 1<sup>st</sup> Delphi |  | 2<sup>nd</sup> Delphi |  | Total |  |
| --- | --- | --- | --- | --- | --- | --- |
|  | n (%) | % | N (%) | % | N (%) | % |
| Total | 215 |  | 104 |  | 319 |  |
| **Stakeholder group (> 1 response allowed)** |  |  |  |  |  |  |
| Stakeholder/policy maker/health services | 9 | 4% | 15 | 9% | 24 | 6% |
| Practitioner/clinician | 110 | 43% | 82 | 50% | 192 | 46% |
| Researcher | 13 | 5% | 39 | 24% | 52 | 12% |
| Patient/client/consumer | 84 | 33% | 21 | 13% | 105 | 25% |
| Not specified | 38 | 15% | 7 | 4% | 45 | 11% |
| **TOTAL (>100% as more than 1 response allowed)** | **254** | **100%** | **164** | **100%** | **418** | **100%** |
| **Geography** |  |  |  |  |  |  |
| Australia | 31 | 15% | 9 | 9% | 40 | 13% |
| Austria | 0 | 0% | 1 | 1% | 1 | 0% |
| Belgium | 2 | 1% | 1 | 1% | 3 | 1% |
| Canada | 15 | 7% | 8 | 8% | 23 | 7% |
| Germany | 1 | 0% | 0 | 0% | 1 | 0% |
| Greece | 1 | 0% | 0 | 0% | 1 | 0% |
| Hong Kong | 0 | 0% | 1 | 1% | 1 | 0% |
| Italy | 0 | 0% | 3 | 3% | 3 | 1% |
| India | 2 | 1% | 0 | 0% | 2 | 1% |
| Indonesia | 1 | 0% | 0 | 0% | 1 | 0% |
| Ireland | 2 | 1% | 0 | 0% | 2 | 1% |
| New Zealand | 25 | 12% | 2 | 2% | 27 | 8% |
| Portugal | 0 | 0% | 1 | 1% | 1 | 0% |
| Sweden | 1 | 0% | 0 | 0% | 1 | 0% |
| Switzerland | 1 | 0% | 0 | 0% | 1 | 0% |
| South Korea | 0 | 0% | 6 | 6% | 6 | 2% |
| United Arab Emirates | 1 | 0% | 0 | 0% | 1 | 0% |
| United Kingdom | 21 | 10% | 17 | 16% | 38 | 12% |
| United States | 56 | 26% | 51 | 49% | 107 | 34% |
| Not specified/missing | 52 | 25% | 4 | 4% | 59 | 18% |
| **TOTAL Geography** | **212** | **100%** | **104** | **100%** | **319** | **100%** |
| **Age category** |  |  |  |  |  |  |
| 18-29 | 27 | 13% | 3 | 3% | 30 | 9% |
| 30-39 | 47 | 22% | 4 | 4% | 51 | 16% |
| 40-49 | 42 | 20% | 25 | 24% | 67 | 21% |
| 50-59 | 38 | 18% | 38 | 37% | 76 | 24% |
| 60 + | 21 | 10% | 26 | 25% | 47 | 15% |
| Not specified | 40 | 19% | 8 | 8% | 48 | 15% |
| **TOTAL Age Category** | **215** | **100%** | **104** | **100%** | **319** | **100%** |
| **Gender** |  |  |  |  |  |  |
| Male | 8 | 4% | 20 | 19% | 28 | 9% |
| Female | 170 | 79% | 75 | 72% | 245 | 77% |
| Non-binary | 9 | 4% | 1 | 1% | 10 | 3% |
| Not specified/missing | 28 | 13% | 8 | 8% | 36 | 11% |
| **TOTAL Gender** | **215** | **100%** | **104** | **100%** | **319** | **100%** |

#### Response rates and attrition

Respondents were recruited through a variety of platforms, from professional connections to public forum groups, providing a broad range of perspectives. Respondents included 192 practitioners, 105 consumers, 52 researchers, 24 policymakers, ensuring inclusion of clinical experts, as well as non-clinical consumers and policy makers, reducing bias from over representation of any of the collaborative groups. We aimed for a sample size of between 20-30 respondents per group, in line with COS development guidelines to reach data saturation (55).

Previous research has acknowledged that attrition between surveys is a common problem. A systematic review demonstrated that surveys that contain a higher number of items have a significantly lower response rate and attrition between surveys (39). In our study, the number of items on each of the five surveys was notably high, which may have impacted the attrition rate, which was on average 52% (ranging from 40% to 68% between surveys) (Table 3). This rate was around the average reported in the review by Gargon (2021). We used strategies such as email reminders, tight timeframes, and countdowns, flyers, and personal approaches for groups, informed by both our research experience and recommendations in the literature (39), which helped to mitigate attrition and improve response rates.

**Table 3.** Attrition for respondents across Delphi surveys.

| Delphi survey condition | Round 1 Respondents (n) | Round 2 Participants (n) | Total Attrition (%) |
| --- | --- | --- | --- |
| Menstruation | 57 | 34 | 40 |
| Pregnancy | 43 | 23 | 46 |
| PCOS | 37 | 19 | 49 |
| Infertility | 37 | 15 | 59 |
| Menopause | 41 | 13 | 68 |

#### Survey results

Overall, survey round one produced 442 outcomes which received an average importance score of ≥ 7 and were carried through to the second round. Between 13 and 117 outcomes were retained in each of the conditions of menstruation (n=85), pregnancy (n=141), PCOS (n=13), fertility (n=117), and menopause (n=86) respectively (Table 4).

These outcomes were included in survey round two, which produced 396 outcomes receiving an average importance score of ≥ 7 in the conditions of menstruation (n=74), pregnancy (n=133), PCOS (n=12), fertility (n=103) and menopause (n=74) respectively. These outcomes were collated by the researchers into six final lists, including a separate acupuncture outcomes list (Table 4), and one each for the five women’s health conditions (Appendix 7). These were brought forward for discussion in the final consensus meeting. The results of Delphi surveys are detailed in Appendix 8.

**Table 4.** Number of outcomes deemed eligible for Delphi round two and final consensus meeting.

| Women's health condition | Outcomes averaging a score of $\geq 7$ (out of 9) in importance (n) | |
| --- | --- | --- |
|  | Round 1 | Round 2 |
| Menstruation | 85 | 74 |
| Pregnancy | 141 | 133 |
| PCOS | 13 | 12 |
| Fertility | 117 | 103 |
| Menopause | 86 | 74 |
| TOTAL | 442 | 396 |

#### Acupuncture treatment specific outcome results

Outcomes from across the five surveys contained similar outcomes for the categories of EAM, safety and feasibility outcomes. There were 31 outcomes from the first survey and 30 from the second. Researchers subsequently formed an outcomes list intended to be primary outcome set commonly collected across all surveys of acupuncture trials in women’s health regardless of condition studies. The individual outcome sets for each of the women’s health conditions are in addition to this list (and reported separately). This list was compiled for discussion at the consensus meeting (Table 5).

**Table 5:** Acupuncture treatment specific outcomes.

|  | Category | Survey 1 outcomes<br>n=31 | Survey 2 outcomes<br>n=30 |
| --- | --- | --- | --- |
| 1 | Acupuncture specific outcomes (EAM) | • EAM system / style / type (e.g., TCM, EAM, Japanese, Korean etc. systems)<br>• EAM diagnosis - diagnostic criteria (e.g., TCM diagnosis)<br>• Perceived changes (patient reported)<br>• Acupuncture protocol used (protocolized or individualized)<br>• Treatment approach<br>• Tongue diagnosis<br>• Pulse diagnosis<br>• Abdominal palpation findings (e.g., pain, temperature, etc.)<br>• Frequency of acupuncture treatment per week / month<br>• Duration/length of treatment (time – mins/hrs.)<br>• Duration of treatment (in weeks)<br>• Relative temperature of 3 Jiaos<br>• Satisfaction with care<br>• Satisfaction - overall (e.g., symptoms, pain management, treatment)<br>• Worsening of symptoms<br>• Person wellbeing<br>• Severe pain, tingling, swelling<br>• Abdominal discomfort<br>• Perceived patient / client experience / acceptability of acupuncture / acupressure / moxibustion / cupping treatment<br>• Quality of life (QoL)<br>• Depression<br>• Low mood<br>• Anxiety<br>• Stress<br>• Social and work impact<br>• Effect on activities of daily living<br>• Severity of condition<br>• Length of condition<br>• Cost effectiveness or cost outcomes | • Patient reported outcome measures (PROMs)<br>• EAM system / style / type (e.g., TCM, EAM, Japanese, Korean, etc. systems)<br>• EAM diagnosis - diagnostic criteria (e.g., TCM diagnosis)<br>• General wellbeing<br>• Perceived changes to symptoms (patient reported changes) with treatment<br>• Satisfaction - overall (e.g., symptoms, pain management, treatment)<br>• Self-reported quality of life (QoL)<br>• Perceived acceptability of acupuncture treatment<br>• Perceived changes (patient reported) with treatment<br>• Satisfaction - overall symptom management<br>• Satisfaction with overall care<br>• Worsening of symptoms<br>• Person wellbeing<br>• Severe pain, tingling, swelling<br>• Abdominal discomfort<br>• Perceived patient / client experience /acceptability of acupuncture/ acupressure / moxibustion / cupping treatment<br>• Quality of life (QoL)<br>• Depression<br>• Low mood<br>• Anxiety<br>• Stress<br>• Social and work impact<br>• Effect on activities of daily living<br>• Severity of condition<br>• Length of condition<br>• Cost effectiveness or cost outcomes<br>• Acceptability of acupuncture treatment<br>• Individual treatment costs<br>• Cost of materials, patient expenses |
| 2 | Safety outcomes | • Minor adverse events - (from needling e.g., bruising, fatigue, pain with needling, fainting, nausea, headache)<br>• Serious adverse events (from needling – e.g., infection, pneumothorax, significant worsening of symptoms) | • Minor adverse events - (from needling e.g., bruising, fatigue, pain with needling, fainting, nausea, headache)<br>• Serious adverse events (from needling – e.g., infection, pneumothorax, significant worsening of symptoms) |

### Consensus meeting

After voting was completed for round two of the surveys, 234 outcomes were compiled into six proposed COS, and brought for discussion at a post-conference consensus workshop after the 2025 SAR conference. Of the outcomes, 202 were outcomes deemed important by Delphi survey respondents, and an additional 32 outcomes were presented from previous related COS. Using a two-round World Cafe design (41), 65 participants from over ten countries took part in the consensus meeting, all of whom were attendees at the SAR 2025 conference. After discussion of the proposed outcomes, participants numbered each outcome in ascending order of importance (1 = most important, 2 = second most important, etc.). Appendix 10 contains details of the outcome rankings for each domain. Rankings from each participant were recorded and averaged, and these averages were used to inform the creation of a final consensus-approved COS for each domain.

#### Acupuncture set of trial reported core outcomes (ASTRO)

In round one of the consensus meeting, participants discussed the proposed COS for acupuncture specific outcomes as a whole group and ranked the outcomes presented, noting down additional comments. All researchers were present to discuss queries or ideas until consensus was reached by participants on the final COS for acupuncture specific outcomes. Outcomes were collated according to rankings and are presented in order in Table 6. The group agreed on a name for the overarching core set; ‘*acupuncture sets of trial reported core outcomes*’ – *ASTRO.* This ASTRO provides a common primary COS, for which each of the women’s health conditions are extensions.

**Table 6.** Final - Acupuncture Sets of Trial Reported Outcomes (ASTRO) Core outcome set (COS)

|  | Acupuncture | Consensus Outcomes | Includes |
| --- | --- | --- | --- |
| 1 | Safety outcomes (Adverse events) | 1. Serious adverse events (SAEs)<br>2. Minor adverse events (AEs) | ● Definition: Any AE* from needling or treatment, which requires medical intervention or jeopardizes ongoing health or wellbeing – e.g., infection, pneumothorax, burns, significant worsening of symptoms)<br>● Any minor side-effects from treatment with needles/moxa/cupping/pressure - e.g. bruising, fatigue, pain with needling, fainting, nausea, dizziness, headache). Transitory adverse events - e.g. passing pain or discomfort with treatment<br>*Lists of SAEs and AEs as defined by Witt *et al.* (2009) (2) and/or Baumler *et al.* (2022) (3). |
| 2 | Acupuncture specific outcomes (EAM) | 1. Primary reported outcome measure (PROM) | ● Perceived changes from treatment in patient reported outcome measure (PROM) for chief complaint. |
|  |  | 2. PROM - (all other outcomes)<br>3. Patient experience / satisfaction<br>4. Demographics<br>5. Mood / cognition<br>6. Sleep<br>7. Pain<br>8. Change in EAM diagnostic indices | Perceived change in PROM for anything other than chief complaint.<br>Satisfaction - overall symptom management, pain management, treatment experience<br>Age, race/ethnicity, sex, gender identity<br>Changes to mood and/or cognition<br>PROM for sleep<br>Quality / experience inventory/scale of pain<br>Change in EAM diagnostic measure – e.g., tongue, pulse, abdominal reflex, etc. |
| 3 | Feasibility | 1. Acceptability<br>2. Cost | Acceptability of acupuncture treatment<br>Satisfaction with overall care<br>Individual treatment costs<br>Patient expenses<br>Cost of materials |
|  | Covered in STRICTA | Acupuncture protocol used<br>Treatment approach<br>Treatment parameters | Protocolized or individualized<br>EAM system / style / type (e.g., TCM, EAM, Japanese, Korean etc. systems)<br>Duration / length of treatment (time)<br>Duration of treatment (weeks)<br>Frequency of treatments per week |

#### STRICTA Guidelines

The group considered outcomes that are already contained in the ‘*STandards for Reporting Interventions in Clinical Trials of Acupuncture’* (STRICTA) guidelines (4). The STRICTA guidelines provide a checklist for improving reporting transparency and acupuncture treatment characteristics to improve the standard of acupuncture protocols and clinical trials. All outcomes contained in STRICTA are already recommended to be included in reporting for acupuncture trials and therefore are not included in the proposed *ASTRO* Women’s Health COS (but were included for participants’ information). A comparison of the role of each ASTRO and STRICTA is provided below. The group was presented with a list of previously published definitions for acupuncture-related adverse events (AEs), and serious adverse events (SAEs) by Witt *et al.,* (2) and Baumler *et al.,* (3). The group adopted these definitions for consistency in reporting of AEs and SAEs.

#### Complementary Roles of ASTRO and STRICTA

Using the comparison below, the complementary nature of ASTRO and STICTA are evident

| Feature | ASTRO COS | STRICTA Guidelines |
| --- | --- | --- |
| Primary focus | Core outcomes to measure and report | Intervention details to describe |
| Developed using COMET methodology? | • Yes | • No |
| Emphasis on East Asian Medicine (EAM) | • Strong - includes diagnostic criteria and context | • Limited to rationale for point selection |
| Includes PROMs and patient experience | • Yes | • No |
| Covers safety and adverse events | • Yes | • No |
| Focus on intervention fidelity | • Not primary | • Yes - detailed protocol description |
| Role in clinical trials | • Defines *what* to measure | • Defines *how* to describe treatment |

Together, ASTRO and STRICTA comprise a thorough and transparent approach to acupuncture trial design and reporting, particularly in complex and patient-centered areas such as women’s health.

#### Women’s Health Conditions COS

Participants were then divided into researcher-facilitated small groups for discussion of each women’s health condition, based on interest and expertise. There was a minimum of two and a maximum of 12 participants in each group. The groups discussed and ranked the outcomes until consensus was reached for a core set for each women’s health condition. These are reported separately for each condition as an ASTRO extension (See Appendix 10).

Where previous condition-specific COS existed, as described previously, these referenced outcomes were listed for consideration as they related to acupuncture trials for treatment of the specific condition. Where relevant, these COS were cited as recommended for inclusion in studies, in addition to our proposed ASTRO-condition specific COS. Studies by Al Wattar *et al.* (45) for PCOS, Herman *et al.* (47), Cooper *et al.* (46) and Kim *et al.* (48) for menstrual health conditions, the Harbin group (51) and Duffy *et al.* (50) for fertility, Remus *et al.* (52), Smith *et al.* (43) and Jones *et al.* (44) for pregnancy conditions, and Lensen *et al.* (49) for menopause COS were considered by our panel to be relevant to our study and selected relevant outcomes were presented. Investigators are therefore encouraged to consider both our COS and the COS from those listed for reporting on these domains as a minimum. These are considered in more detail in the reporting of the ASTRO extension studies.

There was general agreement that acupuncture specific outcomes are important for reporting in all trials in addition to the STRICTA guidelines, and should be extended beyond just those including women’s health conditions. Use of previously defined AEs and SAEs for acupuncture related studies was readily agreed as important and comprehensive in their definition. Importantly, participants reflected that these outcomes should be able to be collected by any clinical practitioner and should be a minimum for inclusion in clinical note taking. This would facilitate the presentation of aggregated data and case histories, as well as personal data tracking.

## Discussion

### Main findings

In this study, we report on the development of the first COS for harmonizing acupuncture research in women’s health to establish the ASTRO framework. The ASTRO framework is informed by the domains of sleep, pain, mood/cognition, and safety/feasibility, which reflect outcomes for whole person health, integral to concepts and philosophies of EAM systems, inclusive of acupuncture studies. This framework provides core outcomes for safety, EAM specific outcomes and feasibility, relevant for all acupuncture studies, with extensions relevant for specific women’s health conditions in addition to the ASTRO Women’s Health COS. This also provides a foundation point for the development of tools to measure the outcomes specified and to established what would be minimally important clinical differences (MICD) to be reported, reflecting the smallest change in an outcome measure that a patient would perceive as beneficial, potentially leading to a change in their management.

The final core set included 12 outcomes categorized into three domains of safety, EAM outcomes and feasibility, which could be readily conducted in a clinical practice setting. We leveraged extensive evidence syntheses on acupuncture trials for women’s health for EAM, safety and feasibility outcomes (structured searches retrieving 326 papers, and reporting 579 separate outcomes [menstrual health (74), fertility (26), PCOS (23), pregnancy (185), and menopause (18) studies/systematic reviews]) from multiple databases to capture the full range of outcomes and engaged a wide multidisciplinary multinational stakeholder panel in a consensus building Delphi and workshop process. Practitioners’, researchers’ and consumers’ input had a pivotal role in the development of this COS, exemplified by focus on specific outcomes capturing whole person health inherent to EAM philosophies.

### Comparison with literature

Outcomes that are of importance to acupuncture studies in women’s health are likely to be relevant to broader studies of acupuncture. In the COMET database, there are six completed trials of acupuncture interventions, which follow the COMET study design. None of these studies have provided specific outcome sets which include acupuncture or broader EAM outcomes. Two other studies which are listed in the COMET database, each following different study designs. These are specifically pregnancy related and investigate the use of EAM modalities. The first study by Betts *et al.* (2014) (43) reports on a two-round Delphi study to establish a consensus on the use of moxibustion for breech version. In this study, participants consisted only of expert practitioners for the development of a moxibustion protocol. The second study is a Cochrane systematic review which reports on 310 trials examining pharmacological and non-pharmacological analgesic methods for pain relief in labour and birth. They report on acupuncture outcomes among others and suggest ‘some evidence’ to support improved pain management and improved satisfaction with pain relief compared with placebo or standard care, with few reported side-effects. The proposed COS from the Cochrane review relates to labour and birth outcomes (maternal, neonatal and cost) (44). We consider outcomes suggested by both of these studies in the ASTRO-Pregnancy COS.

### Significance

This study addresses a longstanding gap in acupuncture research for women’s health by establishing the first harmonized COS by developing the ASTRO framework. Historically, methodological inconsistencies and fragmented outcome reporting have impeded the comparability and clinical relevance of acupuncture trials, particularly as the field expands and care models become more diverse and complex. By standardizing outcome measures, the ASTRO Women’s Health COS enhances methodological rigor and enables meaningful cross-trial comparisons, directly supporting higher quality evidence synthesis and more actionable research findings.

The ASTRO framework is unique in its explicit integration of EAM principles, prioritizing domains such as pain, sleep, mood/cognition, and safety/feasibility. This approach reflects acupuncture’s whole-person philosophy, capturing outcomes relevant to physical, emotional, and cognitive health. The inclusion of these intersectional themes aligns with patient-centered care models and addresses the limitations of reductionist research paradigms that have historically dominated the field.

### Limitations

The limitations of this study should be considered when interpreting the findings. Firstly, our literature search was limited by the exclusion of non-English language studies. Many relevant EAM treatments have been historically studied in non-English speaking populations, and thus our search yielded many papers written in other languages (e.g., Chinese, Japanese, Korean). The exclusion of these studies due to English language inclusion criteria resulted in the omission of some outcomes that may be relevant for non-English speaking populations. Similarly, most participants originated from high-income, English-speaking countries such as the USA, Australia, and the UK.

Participation was also limited to individuals with digital access, potentially excluding those with lower digital literacy or lacking stable internet connection. These factors may have introduced disparities in representation and limited the generalizability of results.

The attrition rate was high across the two Delphi survey rounds (52% on average), likely due to survey fatigue from the extensive list of outcomes. The results of surveys could vary between those who completed the process versus those who did not, potentially introducing bias into the consensus results.

### Utility

Together, the 12 core outcomes identified in three domains of EAM outcomes, safety, and feasibility form a COS that reflects a balance between clinical relevance and the priorities of patients, practitioners, and other stakeholders. Inclusion of both objective clinical endpoints and subjective patient experiences will allow future research to capture extensive data on women’s health that is patient-centered while maintaining scientific rigor. This will allow for discussion of appropriate tools (questionnaires, scales) for the measurement of outcomes, and what would constitute the MICDs for implementation of research findings.

Standardizing these outcomes and forming this ASTRO framework provides a more robust foundation for future clinical trials for acupuncture in women’s health, informing the development of methodology that reflects patient-centered outcomes and is relevant for clinicians. Future research is planned to expand the ASTRO framework, to report other domains of acupuncture trials for women’s health conditions (ASTRO-menstrual health, ASTRO-fertility, ASTRO-PCOS, ASTRO-pregnancy, ASTRO-menopause) and potentially to other fields where outcome heterogeneity limits collection and synthesis of evidence.

## Conclusion

COS can improve research quality, reduce bias, and ensure that outcomes align with the needs of patients and practitioners by their inclusion in research processes. By standardizing core measures, this ASTRO Women’s Health COS addresses significant challenges in women’s health, bridging the gap between research and practical application and ultimately enhancing the well-being of women. Researchers are encouraged to adopt this ASTRO Women’s Health COS in EAM studies to standardize reporting and enable powerful and impactful evidence synthesis.

## Data Availability

All data produced in the present work are contained in the manuscript

## Acknowledgements

The authors acknowledge the support of the Society for Acupuncture Research (SAR), Healthy Seminars (HS), Evidence Based Acupuncture (EBA), the Obstetrical Acupuncture Association (OBAA), and the Maternity Acupuncture Mentoring and Peer Support (MAMPS) program. We would also like to thank all the people who participated in this study.

## Authors’ roles

All authors contributed to the conceptualization, development, conduct, analysis and write up of this study. More emphasis was given by collaborators in various areas, and these are as follows.

## Authors

Kate M Levett (KML), Valentina Buay (VB), Claudia Citkovitz (CC), Kathleen Lumiere (KL), Rosa Schnyer (RS), Sandro Graca (SG), Wen Tu (WT), Debra Betts (DB), Belinda “Beau” Anderson (BA), Lisa Conboy (LC), Lisa Taylor-Swanson (LTS).

Conceptualization: KML, DB, LTS, CC

Protocol writing: KML, DB, LTS, CC

Ethics application: KML, VB, LTS

Search strategy, data extraction: VB, KML, KL, DB, CC, LTS, RS, SG, BA, LC

Data curation: KML, VB, KL, DB, CC, LTS, RS, SG, BA, LC

Formal analysis, KML, CC, VB

Investigation/moderation of workshops and meetings: KML, KL, DB, CC, LTS, RS, SG, BA, LC

Methodology: KML, KL, DB, CC, LTS, RS, SG, BA, LC

Project oversight: KML, LTS, CC

Writing - original draft: KML, VB

Writing – review and editing: KML, LTS, KL, DB, CC, LTS, RS, SG, BA, LC

All authors have read and agreed to the published version of the manuscript.

## Abbreviations

AE: Adverse event
ASTRO: Acupuncture Sets of Trial Reported Outcomes
COMET: Core outcome measures in effectiveness trials
COSMIN: COnsensus-based Standards for the selection of health Measurement INstruments initiative
COS-STAR: Core Outcome Set–STAndards for Reporting
CM: Complementary medicine
COS: Core Outcome Set
EAM: East Asian medicine
MHT: Menopause hormone therapy
PCOS: Polycystic ovarian syndrome
PRISMA: Preferred Reporting Items for Systematic Reviews and Meta-Analyses
PROM: Patient Reported Outcome Measure
QoL: Quality of life
SAE: Serious adverse event
SAR: Society for Acupuncture Research
SPIRIT: Standard Protocol Items: Recommendations for Interventional Trials statement
STRICTA: Standards for Reporting Interventions in Clinical Trials of Acupuncture
TCM: Traditional Chinese medicine
USA: United States of America
WHO: World Health Organization
WPH: Whole person health

