## Supplemental files for "Acupuncture sets of trial reported core outcomes (ASTRO) for women’s health across the lifespan"

### List of Appendices

**Appendix 1 Core Outcome Standards for Reporting - The COS-STAR Statement.**	37

**Appendix 2. Systematic literature Search Strategy**	38

**Appendix 3 Summary of results and outcomes from literature search**	40

**Appendix 4. Outcomes derived from literature review**	40

**Appendix 5	Initial outcomes and rationale for decisions from the research team’s workshops**	40

**Appendix 6. Recruitment strategy**	41

**Appendix 7 Participant details for Round one surveys for each condition**	42

**Appendix 8. Participant details for Round two surveys for each condition**	43

**Appendix 9 Condition subcategories**	43

**Appendix 10 Initial list of outcomes included in surveys**	44

**Appendix 11 Core outcomes from Jones et al. (2012)**	52

**Appendix 12 Delphi survey results and attrition analysis**	52

**Appendix 13 Domain specific outcomes used for consensus surveys**	52

**Appendix 14 Consensus survey rankings**	52

**Appendix 15 Consensus meeting 1 and 2 reports**	52

**Appendix 1 Core Outcome Standards for Reporting - The COS-STAR Statement.**

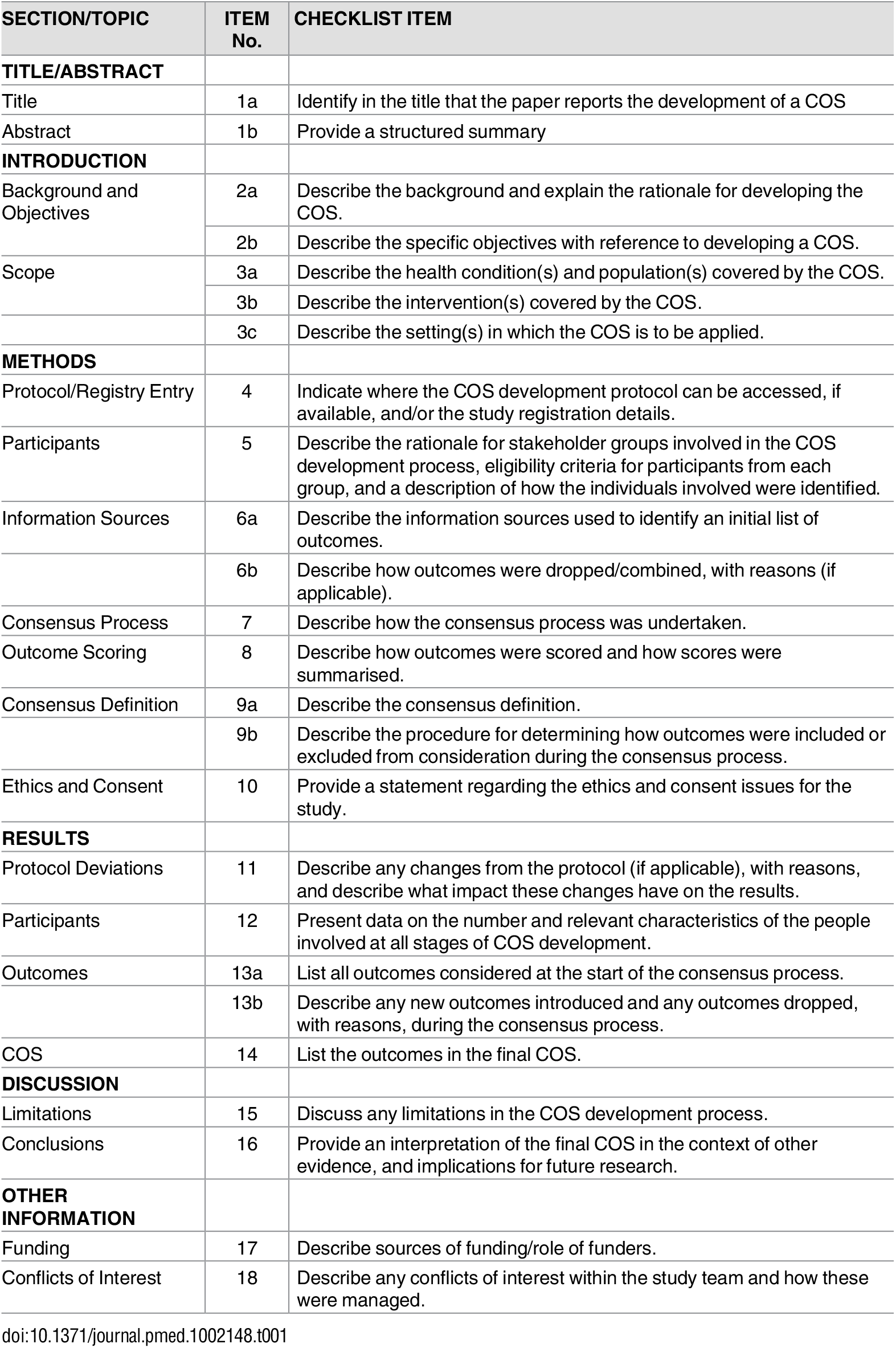

**Appendix 2. Systematic literature Search Strategy**

We note that the following databases use variations in truncations and wildcards to account for variations in spelling and endings of words:

- **Ovid**: truncations use ‘*’ and wildcards are ‘?’, ‘$’ or ‘#’.
- **PubMed**: truncations, suggests using MeSH term, plural forms or synonyms manually, and wildcards are ‘*’.
- **EBSCO:** truncations use ‘*’, wildcards are ‘?’, ‘#’.
- **Wiley**: truncations use ‘*’, and wildcard use ‘?’.

| **1** | T&CM practices and practitioners |
| --- | --- |
| **Search Terms** | Medical Subject Headings (MeSH)  Acupuncture [*methods]; Acupressure [*methods]; Acupuncture Therapy [*methods]; Auricular acupuncture [*methods]; moxibustion [*methods]; Chinese medicine / TCM/ East Asian Medicine / Traditional East Asian Medicine  Keywords CONTAINS "acu*” or “acupoint" or "acupressure" or "acupressure-acupuncture therapy" or "acupuncture" or "electro-acupuncture" or "electro-magnetic" or "electroacupuncture" or "electrical stimulation" or "acupuncture" or "auricular acupressure" or "auricular pressure"or "needle insertion" or "moxibustion" or "Tui Na" or OR “acupotom*” OR “pharmacopuncture” OR “meridians” OR “jing luo” OR “jingluo” OR "Medicine, Chinese Traditional" OR “Chinese medicine” OR “Chinese traditional medicine” OR “East Asian Medicine” OR “EAM” OR “Traditional East Asian Medicine” OR “TEAM”  Title CONTAINS "acupoint" or "acupressure" or "acupressure-acupuncture therapy" or "acupuncture" or "electro-acupuncture" or "electro-magnetic" or "electroacupuncture" or "electrical stimulation" or "moxibustion" or "*acupuncture" or "auricular acupressure" or "auricular pressure" or "needle insertion" or "Tui Na" or “TCM” or “traditional Chinese medicine” or “Chinese medicine” or “pharmacopuncture” or “meridians”  AND |
| **2** | WHOLE PERSON HEALTH  Population: biological females, reproductive females, pregnant females, menopausal females |
| **Search Terms** | MESH: (women) OR (female) OR (reproductive) OR (fertility) OR (gyn$ecology) OR (menstrual health) OR (womens health)  OR |
| **3** | MENSTRUAL HEALTH, WOMENS HEALTH, DYSMENORRHOEA  Gynaecology and Fertility Group; Menstrual disorders and subfertility database search |
| **Search Terms** | MENSTRUAL HEALTH  DYSMEORRHOEA  Keywords CONTAINS "dysmenorrhea" or "Dysmenorrhea-Symptoms" or "dysmenorrhoea" or "pain-dysmenorrhea" or "pain-pelvic" or "pelvic pain" or "menstrual cramps" or "menstrual pain" or "primary dysmenorrhea" or "*Dysmenorrhea"or "menstrual distress” or "*menstrual pain"or "primary dysmenorrhea" or Title CONTAINS "dysmenorrhea" or "Dysmenorrhea-Symptoms" or "dysmenorrhoea" or "pain-dysmenorrhea" or "pain-pelvic" or "pelvic pain" or "menstrual cramps" or "menstrual pain" or "primary dysmenorrhea" or "*Dysmenorrhea" or "menstrual distress" or "*menstrual pain"  OR  "Oligo-amenorrhea" or "oligoamenorrhea" or "oligoanovulatory" or "oligohypomenorrhea" or "amenorrhea" or "amenorrhoea" or "Oligo-amenorrhea" or "oligoamenorrhea" or "oligoanovulatory" or "oligohypomenorrhea" or "amenorrhea" or "amenorrhoea"  PubMed as an example, the search terms and strategies are as follows: (Dysmenorrhea[mesh] OR dysmenorrhea[tiab] OR "Pain, Menstrual"[tiab] OR "Menstrual Pain"[tiab] OR "Menstrual Pains"[tiab] OR "Pains, Menstrual"[tiab] OR "Menstruation, Painful"[tiab] OR "Menstruations, Painful"[tiab] OR "Painful Menstruation"[tiab] OR "Painful Menstruations"[tiab] OR "Primary dysmenorrhea"[tiab]).  PCOS  Keywords CONTAINS "polycystic ovary morphology" or "*Polycystic Ovary Syndrome" or "PCOS" or "hirsutism" or Title CONTAINS "polycystic ovary morphology" or "*Polycystic Ovary Syndrome" or "PCOS" or "hirsutism"  ENDOMETRIOSIS  (“endometriosis” OR “endomet*” OR “pelvic pain” OR “dysmenorrhoea”)  OR |
| **4** | FERTILITY AND INFERTILITY |
|  | FERTILITY  "Subfertility-Female" or "Polycystic Ovary Syndrome" or "PCOS" or "endometriosis" or "subfertility" or "unexplained and endometriosis related infertility" or "unexplained infertility" or "unexplained subfertility" or Title CONTAINS "Polycystic Ovary Syndrome" or "subfertility"  OR |
|  | IVF and ART |
|  | Keywords CONTAINS "ART" or "assisted reproduction" or "assisted reproduction techniques" or "IVF" or "ICSI" or "in vitro fertilisation" or "in-vitro fertilisation techniques" or "in vitro fertilization" or "in vitro maturation" or "intracytoplasmic sperm injection" or "subfertility" or "Infertility" or "IUI" or "Intrauterine Insemination" or "*Embryo Transfer" or "ET" or Title CONTAINS "ART" or "assisted reproduction" or "assisted reproduction techniques" or "IVF" or "ICSI" or "in vitro fertilisation" or "in-vitro fertilisation techniques" or "in vitro fertilization" or "in vitro maturation" or "intracytoplasmic sperm injection" or "subfertility" or "Infertility" or "IUI" or "Intrauterine Insemination" or "*Embryo Transfer" or "ET"  OR |
| **5** | Pregnancy and birth database search |
| **Search Terms** | “labo$r” OR childbirth (ti,kw) OR labo$r (ti,kw) or Childbirth: (ti,kw) or pain* manag* (ti,kw) or Pain* labo*r (ti,kw) or contraction* (ti,kw)  “breech” OR “malposition” OR “cephalic version” OR “breech version” OR "breech presentation"[MeSH Terms] OR ("breech"[All Fields] AND "presentation"  [All Fields]) OR "breech presentation"[All Fields] OR ("labour presentation"[All  Fields] OR "labor presentation"[MeSH Terms] OR ("labor"[All Fields] AND  "presentation"[All Fields]) OR "labor presentation"[All Fields])  OR |
| **6** | Menopause and menopausal transition database search |
| **Search Terms** | "Menopause"[Mesh] OR "Climacteric"[Mesh:NoExp] OR menopause[Text Word] OR  menopaus*[Text Word] OR climacteric[tw]) |

**Appendix 3 Summary of results and outcomes from literature search**

| **Women’s health condition** | **Initial search** | **Duplicates** | **Screening** | **Excluded** | **Records retrieved** | **Excluded** | **Assessed for eligibility** | **Included in review** | **Outcomes** |
| --- | --- | --- | --- | --- | --- | --- | --- | --- | --- |
| **Menstrual health** | 7074 | 814 | 6260 | 4731 | 1529 | 1433 | 96 | 74 | 117 |
| **PCOS** | 13460 | 13366 | 101 | 101 | 56 | 0 | 56 | 26 | 94 |
| **Fertility** | 3525 | 2090 | 1445 | 760 | 685 | 589 | 96 | 23 | 80 |
| **Pregnancy** | 2766 | 2284 | 490 | 207 | 283 | 19 | 264 | 185 | 171 |
| **Menopause** | 4124 | 450 | 3674 | 2873 | 801 | 705 | 96 | 18 | 117 |
| **TOTAL** | 30949 | 19004 | 11970 | 8672 | 3354 | 2746 | 608 | 326 | 579 |

**Appendix 4. Outcomes derived from literature review**

Due to space constraints, Appendix 4 containing the full list of outcomes from the literature review is available from the author upon request.

**Appendix 5	Initial outcomes and rationale for decisions from the research team’s workshops**

Due to space constraints, Appendix 5 containing initial outcomes and rationale for decisions from the research team’s workshops is available from the author upon request.

**Appendix 6. Recruitment strategy**

| Condition | Platform / Organisations used for recruitment |
| --- | --- |
| All conditions | - Australian Acupuncture and Chinese Medicine Association (AACMA) - Australian Traditional Medicine Society (ATMS) - American Society of Acupuncturists (ASA) - Philadelphia College of Osteopathic Medicine (PCOM) - Maternity Acupuncture Mentoring and Peer Support group (MAMPS) - Subreddit: R/ChineseMedicine - Acupuncture New Zealand conference |
| Pregnancy | - Maternity Acupuncture Mentoring and Peer Support group (MAMPS) - Midwives participating in acupuncture course - **Subreddits:**   - R/Pregnancy Problems   - R/Gestational Diabetes   - R/Science Based Parenting   - R/BB30: Pregnancies in women over 30 |
| Polycystic Ovarian Syndrome | - **Subreddits:**   - R/PCOSloseit: PCOS weight management   - R/PCOS Management   - R/TTCPCOS: Trying to have a baby with PCOS |
| Menstrual Health | - Subreddit: R/Periods |
| Fertility | - Maternity Acupuncture Mentoring and Peer Support group (MAMPS) |
| Menopause | - Australian Acupuncture and Chinese Medicine Association (AACMA) - Australian Traditional Medicine Society (ATMS) - American Society of Acupuncturists (ASA) - Subreddit: R/ChineseMedicine - Acupuncture New Zealand conference |

**Appendix 7 Participant details for Round one surveys for each condition**

|  | Condition | Responses | Gender | Age | Country | Role (>1 response allowed) | Outcomes (n) |
| --- | --- | --- | --- | --- | --- | --- | --- |
| 1 | Menstrual health | 57 | F: 49  M: 2  N/B: 2  Missing: 4 | 18-29: 16  30-39: 17  40-49: 6  50-59: 6  60+: 3  Missing: 19 | Aust/NZ: 7  Belgium: 2  USA: 18  Canada: 5  UK/Ireland: 16  India: 2  Missing: 4 | Practitioner: 21  Researcher: 4  Patient/consumer: 29  Policy maker: 1  Missing: 6 | 60 |
| 2 | PCOS | 37 | F: 28  M: 2  N/B: 3  Missing: 4 | 18-29: 5  30-39: 8  40-49: 4  50-59: 4  60+: 5  Missing: 11 | Aust: 3  NZ: 8  Germany: 1  USA: 7  Canada: 1  UK/Ireland: 1  Indonesia: 1  Missing: 11 | Practitioner: 13  Researcher:  Patient/consumer: 12  Policy maker:  Missing: 12 | 46 |
| 3 | Fertility | 37 | F: 28  M: 2  N/B: 3  Missing: 4 | 18-29: 2  30-39: 9  40-49: 10  50-59: 9  60+: 3  Missing: 4 | Aust: 6  NZ: 4  Switzerland: 1  Sweden: 1  USA: 14  Canada: 0  UK/Ireland: 1  Missing: 10 | Practitioner:24  Researcher: 3  Patient/consumer: 13  Policy maker: 1  Missing: 4 | 27 |
| 4 | Pregnancy | 43 | F: 34  M: 0  N/B: 0  Missing: 9 | 18-29: 4  30-39: 11  40-49: 11  50-59: 6  60+: 2  Missing: 9 | Aust: 9  NZ: 7  USA: 9  Canada: 3  UK/Ireland: 1  Cyprus: 1  Missing: 13 | Practitioner: 25  Researcher: 2  Patient/consumer: 12  Policy maker: 1  Missing: 9 | 16 |
| 5 | Menopause | 41 | F: 31  M: 2  N/B: 1  Missing: 7 | 18-29: 0  30-39: 2  40-49: 11  50-59: 13  60+: 8  Missing: 7 | Aust: 6  NZ: 4  Switzerland:  USA: 8  Canada:6  UK/Ireland: 2  United Arab Emirates: 1  Missing: 14 | Practitioner: 27  Researcher: 4  Patient/consumer: 18  Policy maker: 6  Missing: 7 | 52 |
|  | Total | 215 | F: 170 M: 8 N/B: 9 Missing: 28 | 18-29: 27  30-39: 47  40-49: 42  50-59: 38  60+: 21  Missing: 50 |  | Practitioner: 110  Researcher: 13  Patient/consumer: 84  Policy maker: 9  Missing: 38 |  |

**Appendix 8. Participant details for Round two surveys for each condition**

|  | Condition | Responses | Gender | Age | Country | Role (>1 response allowed) | Outcomes (n) |
| --- | --- | --- | --- | --- | --- | --- | --- |
| 1 | Menstrual health | 34 | F: 22  M: 9  N/B: 0  Missing: 3 | 18-29: 2  30-39: 2  40-49: 9  50-59: 10  60+: 8  Missing: 3 | Aust: 6  NZ: 0  Belgium: 1  USA: 16  Canada: 1  UK/Ireland: 16  Italy: 2  Hong Kong: 1  South Korea: 1  Portugal: 1  Missing: 4 | Practitioner: 23  Researcher: 9  Patient/consumer: 5  Policy maker: 1  Missing: 3 | 74 |
| 2 | PCOS | 19 | F: 16  M: 2  N/B: 1  Missing: 0 | 18-29: 0  30-39: 0  40-49: 6  50-59: 9  60+: 4  Missing: | Aust: 2  USA: 13  Canada: 1  UK/Ireland:  Italy: 1  Austria: 1  South Korea: 1  Missing: 0 | Practitioner: 16  Researcher: 7  Patient/consumer: 4  Policy maker: 4  Missing: 0 | 12 |
| 3 | Fertility | 15 | F: 12  M: 2  N/B: 0  Missing: 1 | 18-29: 0  30-39: 0  40-49: 3  50-59: 6  60+: 5  Missing: 1 | Aust: 0  NZ: 0  South Korea: 1  USA: 8  Canada: 3  UK/Ireland: 1  Missing: 2 | Practitioner: 13  Researcher: 4  Patient/consumer: 3  Policy maker: 2  Missing: 1 | 103 |
| 4 | Pregnancy | 23 | F: 16  M: 3  N/B: 0  Missing: | 18-29: 1  30-39: 2  40-49: 3  50-59: 9  60+: 4  Missing: 4 | Aust: 0  NZ: 2  USA: 11  South Korea: 2  Canada: 1  Missing: 7 | Practitioner: 18  Researcher: 12  Patient/consumer: 4  Policy maker: 4  Missing: 3 | 133 |
| 5 | Menopause | 13 | F: 9  M: 4  N/B: 0  Missing: 0 | 18-29: 0  30-39: 0  40-49: 4  50-59: 4  60+: 5  Missing: 0 | Aust: 1  NZ: 0  Sout Korea: 1  USA: 3  Canada: 2  UK/Ireland: 0  Missing: 5 | Practitioner: 12  Researcher: 7  Patient/consumer: 5  Policy maker: 4  Missing: 0 | 74 |
|  | Total | 104 | F: 75  M: 20  N/B: 1  Missing: 4 | 18-29: 3  30-39: 4  40-49: 25  50-59: 38  60+: 26  Missing: 8 |  | Practitioner: 82  Researcher: 39  Patient/consumer: 21  Policy maker: 15  Missing: 7 |  |

**Appendix 9 Condition subcategories**

| Condition | Subcategories | Number of subcategories (n) |
| --- | --- | --- |
| Reproductive health and endometriosis | Patient experience  Pain  Sleep  Mood/ Cognition  Endometriosis  Safety and adverse events  Acupuncture/EAM specific outcomes  Menstrual health and history  Other | 9 |
| Fertility | Patient experience  Pain  Sleep  Mood  Cognition  Safety Outcomes  Acupuncture or EAM related outcomes  General fertility  Fertility markers  Fertility pregnancy outcomes | 10 |
| PCOS | Patient experience  Pain  Sleep  Mood/ Cognition  PCOS specific outcomes  Anthropomorphic  Sexual and Menstrual Health History  PCOS treatment and safety  Acupuncture & Safety  Other | 10 |
| Pregnancy | Maternal experience  Pain  Sleep  Mood  Lower back and pelvic pain  Nausea and vomiting in pregnancy (NVP), Hyperemesis Gravidarum (HG)  Breech or Malpresentation  Preeclampsia  Labour preparation  Perinatal  Postnatal  Safety outcomes  Acupuncture or EAM specific safety outcomes  Other | 14 |
| Menopause | Patient experience  Pain  Sleep  Mood  Cognition  Sexual & urogenital  Safety, adverse events  Acupuncture or EAM related outcomes  Other | 9 |

**Appendix 10 Initial list of outcomes included in surveys**

| Reproductive health and endometriosis | | |
| --- | --- | --- |
| Patient experience | Pain | Sleep |
| Perceived experience/acceptability of acupuncture treatments  Participant/patient demographics (e.g., age, ethnicity, etc.)  Social determinants of health (socio-economic status, geographic location)  General wellbeing  Perceived changes to symptoms (patient reported changes) with treatment  Satisfaction – overall (e.g. symptom, pain management, treatment)  Self-reported QoL  Satisfaction with care  Menstrual health | Pain condition  Quality of life  Severity of pain  Duration of pain  Profile of pain  Type of pain  Disability resulting from pain conditions  Lifespan of pain  Painkillers / pain relief required  Frequency of painkillers taken  Amount of painkillers taken  Seeking help for pain  Pain quality (menstruation)  Pain during sex (dyspareunia)  Pain during urination and defecation  Medication review - full MEQ (morphine equivalent dose)  Non-morphine medications | Insomnia (trouble falling asleep, staying asleep, waking earlier than planned)  Quality of sleep  Hours of sleep  Disruption to sleep  Quality of life (related to sleep)  Severity of condition - sleep and severity rating  Feel refreshed on waking  Sleep behaviors (caffeine, alcohol before sleep)  Sleep medication use |
| Mood/Cognition | Safety Outcomes | EAM or Acupuncture Specific Outcomes (ACU-SOS) |
| Quality of life (QoL)  Depression  Low mood  Anxiety  Stress  Fear  Irritability  Social and work impact  Effect on activities of daily living  Severity of condition  Length of condition  Changes to cognition (e.g. foggy brain)  Self-esteem  Dysphoria (PMDD) | Worsening of symptoms  Person wellbeing  Menstrual outcomes/complications  Minor adverse events - (related to gynecological outcomes, e.g. vomiting, headaches, related to medications)  Serious adverse events (e.g. – gynecological outcomes, e.g. requirement for medical review, hospitalization) | EAM or Acupuncture Specific Outcomes (ACU-SOS)  EAM system / style / type (e.g. TCM, EAM, Japanese, Korean etc. systems)  EAM diagnosis - diagnostic criteria (e.g. TCM diagnosis)  Acupuncture protocol used (protocolized or individualized) Acupuncture protocol  Treatment approach  Tongue diagnosis  Pulse diagnosis  Abdominal palpation findings (e.g. reflexes, pressure pain, temperature etc.)  Frequency of acupuncture treatment per week / month  Duration/length of treatment (time – mins/hrs.)  Duration of treatment (in weeks)  Relative temperature of 3 Jiaos  Cost effectiveness or cost outcomes  Minor adverse events from needles/moxa - (e.g.) bruising, fatigue, pain with needling, fainting, nausea, headache adverse events from needle  Serious adverse events (e.g. – hyperstimulation, infection, pneumothorax) |
| Menstrual health and history | Other | |
| General Fertility  Lifespan of condition  Menstruation history  Menstrual regularity  Basal body temperature  Family history  Amount of blood flow in period  Quality of blood  Length of flow during period  Colour of flow/blood  Age beginning of menses  Symptoms of pain and cycle pattern | Perception of normal cycle  Acne  Anemia  Self-image (self-reported)  Nausea  Vit D  STIs  Meds  Abdominal palpation findings  Days off work  Patient perception of 'normal' menstrual experience  Microbiome  Zinc  Pain correlation with vulvodynia  Fibromyalgia  UTI  Social determinants of health  Birth control  PMDD  Chromium/Calcium  Age of onset | Pain and temperature  Patient experience  Hormonal therapy (Oestroprogestinic)  Pain correlation with MRI and profile of endometriosis  NK cells  Micronutrient profiles  Medication review - full MEQ (morphine equivalent dose)  Non-morphine medications  Infection  Heart rate or pulse rate  Abdominal distention  Tubal patency  HbA1c  Hormones - prescribed supplements  CBC basic blood lab tests  Fatigue  Type of menstrual gear used (e.g. cup, pad, tampons)  History of trauma |
| Fertility | | |
| Patient Experience | Pain | Sleep |
| Perceived acceptability of acupuncture treatment  Participant/patient demographics (e.g., age, ethnicity, etc.)  Social determinants of health (socio-economic status, geographic location)  General wellbeing  Perceived changes (patient reported changes) with treatment  Satisfaction – overall symptom management  Self-reported QoL  Satisfaction with care  Fertility experience | Pain condition Quality of life Severity of pain Duration of pain Profile of pain Type of pain Disability resulting from pain conditions Lifespan of pain Painkillers / pain relief required Frequency of painkillers taken Amount of painkillers taken Seeking help for pain Abdominal distention Pain from treatment (fertility related medication or procedures) | Insomnia (trouble falling asleep, staying asleep, waking earlier than planned)  Quality of sleep  Hours of sleep  Disruption to sleep  Quality of life (related to sleep)  Severity of condition - sleep and severity rating  Feel refreshed on waking  Fatigue  Sleep behaviors (caffeine, alcohol before sleep)  Sleep medication use |
| Mood/Cognition | Safety Outcomes | EAM or Acupuncture Specific Outcomes (ACU-SOS) |
| Quality of life  Depression  Low mood  Anxiety  Perceived stress  Fear  Irritability  Social and work impact  Effect on activities of daily living  Severity of condition  Length of condition  Changes to cognition (e.g. foggy brain, dysphoria)  Self-esteem  Coping  Emotional regulation | Worsening of symptoms  Person wellbeing  Menstrual outcomes  Abdominal discomfort  Early pregnancy loss  Minor adverse events (related to fertility outcomes, e.g. vomiting, headaches)  Serious adverse events (related to fertility outcomes, e.g. requirement for medical review, hospitalization) | Diagnostic criteria (e.g. TCM, EAM, Japanese, Korean systems)  EAM diagnosis - diagnostic criteria (e.g. TCM diagnosis)  Acupuncture protocol used (protocolized or individualized)  Treatment approach  Tongue diagnosis  Pulse diagnosis  Abdominal palpation findings (e.g. reflexes, pressure pain, temperature)  Frequency of treatment (acupuncture) per week  Duration/length of treatment (time)  Duration of treatment (in weeks)  Relative temperature of 3 Jiaos  Cost effectiveness or cost outcomes  Minor adverse events from needles/moxa (e.g. bruising, fatigue, pain with needling, fainting, nausea, headache)  Serious adverse events (e.g. hyperstimulation, infection, pneumothorax) |
| General Fertility | Fertility Markers | |
| Lifespan of fertility issues  Menstruation history  Menstrual regularity  Basal body temperature  Family history  Amount of blood in menstruation  Quality of menstruation  Length of cycle / menstruation | Inflammatory markers  General metabolism  Insulin resistance  Anti-mullerian hormone level (AMH)  LH + FSH levels  Physiological markers  Sex hormones  Menstruation  Progesterone  DHEA  Estradiol Level  Gonadotropin (Gn) dosage and duration  Stress hormones | Testosterone (T)  Thyroid  LDL-C, HDL-C  Women's feelings about self  Antral Follicle Count (AFC)  Endometrial factors  Sex hormone binding globulin (SHBG)  Radial line of endometrial cyst of ovary  17-α-hydroxyprogesterone (17-α-OHP)  Androstenedione (A2)  Metabolites in follicular fluid  OGTT  BP |
| Fertility Pregnancy Outcomes | | Pregnancy |
| Live baby rate  Pregnancy loss rates  Pregnancy loss  Antral follicle count (AFC)  Conception rate  Endometrial receptivity  Follicular development  Fertilization rates  High quality embryo rate  Oocytes/zygotes  Ovarian morphology  Ovulation rate  Natural pregnancy | Transplantable embryo rate  Embryo rate  Retrieved  Morphology  Ovulation rate  Pregnancy confirmed  Chemical pregnancy  Implantation  Rounds of IVF  IUI  Type of IVF  Hormonal stimulation | Collection rates  Genetic conditions  Follicular growth to blastocyst rate  Age of embryos  Sperm quality  Demographics  Health experiences  Safety outcomes |
| Polycystic **Ovarian Syndrome (PCOS)** | | |
| Patient Experience | General Pain | Sleep |
| Perceived experience/acceptability of acupuncture treatments  Participant/patient demographics (e.g., age, ethnicity, etc.)  Social determinants of health (socio-economic status, geographic location)  General wellbeing  Perceived changes to symptoms (patient reported changes) with treatment  Satisfaction – overall symptom management  Self-reported QoL  Satisfaction with care  PCOS specific changes | Pain condition  Quality of life  Severity of pain  Duration of pain  Profile of pain  Type of pain  Disability resulting from pain conditions  Lifespan of pain  Painkillers / pain relief required  Frequency of painkillers taken  Amount of painkillers taken  Seeking help for pain  Pain quality (e.g. ovulation/menstruation)  Abdominal distention  Pain condition (location, type, origin) | Insomnia (trouble falling asleep, staying asleep, waking earlier than planned)  Quality of sleep  Hours of sleep  Disruption to sleep  Quality of life (related to sleep)  Severity of condition - sleep and severity rating  Feel refreshed on waking  Sleep behaviors (caffeine, alcohol before sleep)  Sleep medication use |
| Mood/Cognition | Safety Outcomes | EAM or Acupuncture Specific Outcomes (ACU-SOS) |
| Quality of life (QoL)  Depression  Low mood  Anxiety  Stress  Fear  Irritability  Social and work impact  Effect on activities of daily living  Severity of condition  Length of condition  Self-esteem  Dysphoria (e.g. PMDD) | PCOS complications  Hyperstimulation of ovaries/follicles  Cycle changes  Minor adverse events - related to PCOS outcomes, e.g. vomiting, headaches, related to medications  Serious adverse events - related to PCOS outcomes, e.g. requirement for medical review, hospitalization | EAM system / style / type (e.g. TCM, EAM, Japanese, Korean etc. systems)  EAM diagnosis - diagnostic criteria (e.g. TCM diagnosis)  Acupuncture protocol used (protocolized or individualized)  Treatment approach  Tongue diagnosis  Pulse diagnosis  Abdominal palpation findings (e.g. reflexes, pressure pain, temperature etc.)  Frequency of treatment (acup.) per week  Duration/length of treatment (time)  Duration of treatment (in weeks)  Relative temperature of three jiaos  Cost effectiveness or cost outcomes  Minor adverse events from needles/moxa - (e.g.) bruising, fatigue, pain with needling, feeling faint, clammy, fainting, nausea, headache  Serious adverse events (e.g. hyperstimulation, infection, pneumothorax) |
| Physiological markers | | Anthropomorphic |
| General metabolism  Metabolic markers  Insulin resistance / sensitivity / glucose tolerance  HbA1c  Inflammation markers  LH + FSH (Luteinizing Hormone and Follicle Stimulating Hormone)  Progesterone  17-α-hydroxyprogesterone (17-α-OHP)  Stress hormones (e.g. cortisol) | Androgens / androstenedione (A2)  Free androgen index (FAI)  Presence of cysts (on ultrasound)  Antral follicle count (AFC)  Sex hormone binding globulin (SHBG)  Gonadotropin (Gn) dosage and duration  LDL and HDL-Cholesterol  Thyroid function (e.g. TSH, T3, T4, antithyroid antibodies)  Anemia  Vit D  Blood pressure  Acne  Hirsutism  Length of menstrual cycle (i.e. interval between menstrual cycles)  NK (natural killer) cells  CBC basic blood lab tests | General anthropometric  Fat to muscle ratio  BMI  Waist-to-hip ratio (WHR)  Weight  Diet assessment  Activity level  Physical exercise assessment |
| Sexual/menstrual health history | | Other |
| Birth control taken (type, history)  Amenorrhea (incidence, duration)  Infertility (incidence, duration)  Requirement for assisted reproduction technology (ART) (e.g. IVF, IUI, hormone triggers etc.)  History or presence of STIs (sexually transmitted infections)  History or presence of UTIs (urinary tract infections) | PMS (premenstrual syndrome)  PMS (premenstrual  PMDD (premenstrual dysphoric disorder)  Dysmenorrhea / pelvic pain  Pain correlation with vulvodynia  Sexual dysfunction - pain during sex (dyspareunia)  Type of menstrual gear used (e.g. cup, pad, tampons)  Tubal patency  Pregnancy rates  Infertility | Perception of normal cycle  Age of onset - menarche  Self-image (self-reported)  History of trauma  Psycho-social (e.g. ACES)  Days off work  Sleep quality  Nausea  Microbiome  Medications taken  Hormonal therapy (Oestroprogestinic)  Hormone - prescribed supplements  Micronutrient profiles  Chromium/Calcium  Zinc |
| Pregnancy | | |
| Patient Experience | Pain | Sleep |
| Perceived experience/acceptability of acupuncture treatments  Participant/patient demographics (e.g., age, ethnicity, gestational age, gravidity, parity etc.)  Social determinants of health (socio-economic status, geographic location)  General wellbeing  Perceived changes to symptoms (patient reported changes) with treatment  Satisfaction – overall (e.g. pregnancy, birth, symptom, pain management, treatment etc.)  Self-reported QoL  Satisfaction with care  Fetal movements  Birth experience | Pain condition  Quality of life  Severity of pain  Duration of pain  Profile of pain  Type of pain  Disability resulting from pain conditions  Lifespan of pain  Painkillers / pain relief required  Frequency of painkillers taken  Amount of painkillers taken  Seeking help for pain  Labour pain evaluation (severity, timepoints) | Insomnia (trouble falling asleep, staying asleep, waking earlier than planned)  Quality of sleep  Hours of sleep  Disruption to sleep  Quality of life related to sleep  Severity of condition – sleep and severity rating  Feel refreshed on waking  Sleep behaviors (caffeine, alcohol before sleep)  Sleep medication use |
| Mood/cognition | Safety outcomes | EAM or Acupuncture Specific Outcomes (ACU-SOS) |
| Quality of life (QoL)  Depression  Low mood  Anxiety  Stress  Irritability  Social and work impact  Effect on activities of daily living  Severity of condition  Length of condition  Changes to cognition (e.g. foggy brain)  Self-esteem  Pregnancy/birth fear (e.g. tokophobia) | Worsening of symptoms  Person wellbeing  Pregnancy complications  Preterm contractions  Preterm birth  Maternal and neonatal wellbeing  Labour and birth outcomes  Neonatal outcomes  Admission or extension to hospital stay (maternity, SCN, NICU)  Minor adverse events – (labour and birth related, e.g. cannula site pain, epidural related pain at site)  Serious adverse events (pregnancy and birth related – e.g. admission or extension to hospital stay, epidural injury) | EAM system / style / type (e.g. TCM, EAM, Japanese, Korean etc. systems)  EAM diagnosis - diagnostic criteria (e.g. TCM diagnosis)  Acupuncture protocol used (protocolized or individualized)  Treatment approach  Tongue diagnosis  Pulse diagnosis  Abdominal palpation findings (e.g. pain, temperature etc.)  Frequency of acupuncture treatment per week / month  Duration/length of treatment (time – mins/hrs.)  Duration of treatment (in weeks)  Relative temperature of 3 Jiaos  Cost effectiveness or cost outcomes  Minor adverse events - (from needling e.g.) bruising, fatigue, pain with needling, fainting, nausea, headache  Serious adverse events (from needling – e.g. preterm contractions, preterm birth, infection, pneumothorax) |
| Back and pelvic pain | Breech presentation, posterior or malpresentation | Nausea, vomiting and **hyperemesis in pregnancy** |
| Pain severity  Pain intensity  Length of pain  Gestational age at onset  Duration of pain  Analgesic consumption  Anxiety caused by lower back pain  Depression caused by lower back pain  Disability rating  Clinical exam  Sleep disruption as it relates to pelvic and back pain  Intercostal pain  Sciatica  Hydro-nephrosis in pregnancy  Quality of life (as it relates to back/pubic pain) | Cephalic version at time of intervention – 14 days from treatment onset, or prior to 37 weeks  Duration and frequency of treatment – per day, per week, per course of treatment  Length of treatment (moxa/acupuncture) – days  Need for manual external cephalic version (ECV)  Oligo/polyhydramnios  Anemia or iron deficiency  Position/presentation of baby at birth  Mode of birth – rate of vaginal birth / caesarean section / instrumental or assisted vaginal birth  Manual version success rate  Version following both moxa and ECV  Position of baby – other time points  Gestational age at birth  Pelvic diameter  Safety outcomes  Cord prolapse  Fetal heart rate tracing – pre/post acupuncture, acupressure, or moxa  Placental abruption  Preterm contractions  Baby admission – special care nursery (SCN) or neonatal intensive care unit (NICU)  Length of stay in hospital | Severity of nausea  Severity of vomiting  Duration of nausea  Duration of vomiting  Gestational age at onset of NVP  Current gestational age  Patient-reported changes with treatment  Anxiety caused by nausea/vomiting  Depression caused by nausea/vomiting  Fluid administration  Medications required/taken  Hospital admission  Length of stay in hospital  Presentation/utilization of day stay units or hospital in the home services  Ketones  Number of admissions  Medical / allied health appointments require  Cost assessment  Patient experience with NVP / HG  Postpartum wellbeing  Quality of life  Weight gain/loss  Fluids – 1st trimester  Glucose levels  Diabetes |
| Perinatal outcomes | Postnatal outcomes | Pre-eclampsia |
| Preterm birth <37 weeks’ gestation  Stillbirth >20 weeks' gestation  Neonatal outcomes – gestational age at birth, admissions or observation in SCN or NICU, cord pH, heart rate tracing, ECG, EEG  Onset of labour – spontaneous, induced (IoL, ROM, PGE), none (planned caesarean section)  Mode of birth – normal vaginal birth, instrumental vaginal birth, emergency caesarean section, planned caesarean section  Retained placenta  Uterine involution  Pharmacological medication – epidural, spinal, parenteral analgesia, nitrous oxide, opioid  Non-pharmacological – acupressure/puncture, relaxation, manual therapies, aromatherapy, hypnosis, upright positions etc.  Pain, satisfaction with pain management | Pain / disability  Maternal morbidity  Maternal mortality  Perineal tearing – 1st–2nd degree, 3rd–4th degree, episiotomy  Blood loss and postpartum hemorrhage (PPH) – 1500 mL, >3000 mL  Lactation – breastfeeding within first 2 hours, other lactation outcomes  Infection, uterine involution  Uterine tears, rupture, prolapse  Placental delivery – spontaneous, with synthetic oxytocin  Postpartum depression  Postpartum mortality (up to 12 months)  Quality of life (postpartum)  Mothering experience  Postpartum wellbeing | BP pre/post acupuncture  Labs - preeclampsia panel  Headaches  History of headaches prior to pregnancy   Post-covid preeclampsia rate  Blood panels e.g. platelet count |
| Birth preparation and labour and birth outcomes | | Other |
| Assessment / treatment e.g. cervical ripening, Bishop’s score etc.  Population demographics  Onset of labour spontaneous/induced/no labour (CS)  Gestational age at onset of labour  Medical risk status e.g. induction | Timing of onset  Length of labour  Pain meds used pharmacological, opioid, epidural  Non-pharmacological pain management  Other interventions  Attendance at childbirth and parenting classes  Congenital abnormalities  Place of birth  Model of care | Bell's Palsy  Dyspepsia  Gestational Diabetes  Anemia / Iron deficiency  Socio-economic background  Diabetes in later life following GDM  Constipation |
| Menopause |  |  |
| Patient Experience | Pain | Sleep |
| Perceived acceptability of acupuncture treatment  Participant/patient demographics, e.g., age, ethnicity, etc.  Social determinants of health, socio-economic status, geographic location  General wellbeing  Perceived changes, patient reported changes with treatment  Satisfaction – overall symptom management  Self-reported QoL  Satisfaction with care  Menopause specific changes | Pain condition  Disability resulting from pain conditions  Duration of pain (episodes)  Profile of pain  Pain condition  Quality of life  Severity of pain  Duration of pain  Profile of pain  Type of pain  Disability resulting from pain conditions  Lifespan of pain  Painkillers / pain relief required  Frequency of painkillers taken  Amount of painkillers taken  Seeking help for pain  Bone pain  Headaches  Joint aches  Low back pain  Muscle aches  Neuropathic pain  Vulvar pain  Types of pain  Interference of activities  Fibromyalgia | Insomnia (trouble falling asleep, staying asleep, waking earlier than planned)  Quality of sleep  Hours of sleep  Disruption to sleep  Quality of life related to sleep  Severity of condition - sleep and severity rating  Feel refreshed on waking  Daytime nap  Fatigue  Sleep behaviors (caffeine, alcohol before sleep)  Sleep medication use |
| Mood | Cognition | Safety, Adverse events |
| Quality of life (QoL)  Depression  Low mood  Anxiety  Perceived stress  Fear  Irritability  Social and work impact  Effect on activities of daily living  Severity of condition  Length of condition  Self-esteem  Dysphoria (e.g. PMDD)  Attitude towards menopause  Loss of self-esteem  Coping  Emotional regulation  Impatience  Adverse Childhood Events (ACEs)  Attitudes towards aging | Brain fog  Difficulty concentrating  Memory problems including word recall  More clumsy than usual | Worsening of symptoms  Person wellbeing  Menstrual outcomes  Abdominal discomfort  Hematoma  Hot, skin sensitivity  Nervousness  Severe pain, tingling, swelling  Tremors  Agitation  Fatigue  Dizziness  Minor adverse events related to PCOS outcomes, e.g. vomiting, headaches, related to medications  Serious adverse events related to PCOS outcomes, e.g. requirement for medical review, hospitalization |
| EAM or acupuncture specific outcomes (ACU-SOS) | Sexual and urogenital | Other |
| EAM system / style / type (e.g. TCM, EAM, Japanese, Korean etc. systems)  EAM diagnosis - diagnostic criteria (e.g. TCM diagnosis)  Acupuncture protocol used (protocolized or individualized)  Treatment approach  Tongue diagnosis  Pulse diagnosis  Abdominal palpation findings (e.g. reflexes, pressure pain, temperature)  Frequency of treatment (acupuncture) per week  Duration/length of treatment (time)  Duration of treatment (in weeks)  Relative temperature of 3 Jiaos  Cost effectiveness or cost outcomes  Knowledge of acupuncture safety  Minor adverse events from needles/moxa (e.g. bruising, fatigue, pain with needling, feeling faint, clammy, fainting, nausea, headache, thermoregulation worsening)  Serious adverse events (e.g. infection, pneumothorax) | General urogenital  Change to libido  Urethral (urinary tract) changes  Prone to urinary tract infections  Vaginal dryness  Genital bleeding  Genital skin thinning/changes  General sexual  Sexual satisfaction (including orgasm)  Painful sex (dyspareunia) | Feeling abnormal  Premature menopause before age 40  Feeling abnormal, e.g. ‘I just don’t feel like myself’  General skin changes  General skin dryness  Breast pain or tenderness  Hormone levels  Osteoporosis or osteopenia  Micronutrient levels  Vitamin D levels  Skin dryness  Nose bleeds  Running nose (pre-menopause) with warm foods  Attitudes towards ageing  Work impacts  Experience of own health  Becoming an elder  Breast pain or tenderness |

**Appendix 11 Core outcomes from Jones et al. (2012)**

The following list of core outcomes was developed in collaboration with members of the Pregnancy and Childbirth Group (PCG) consumers’ group:

| Effects of interventions | Safety of interventions | Other outcomes |
| --- | --- | --- |
| Pain intensity (as defined by trialists)  Satisfaction with pain relief (as defined by trialists)  Sense of control in labor (as defined by trialists)  Satisfaction with childbirth experience (as defined by trialists) | Effect (negative) on mother/baby interaction  Breastfeeding (at specified time points)  Assisted vaginal birth  Caesarean section  Adverse effects (for women and infants; review specific)  Admission to special care baby unit/neonatal intensive care unit (as defined by trialists)  Apgar score less than seven at five minutes  Poor infant outcomes at long‐term follow‐up (as defined by trialists) | Cost (as defined by trialists) |

**Appendix 12 Delphi survey results and attrition analysis**

Due to space constraints, Appendix 10 containing the full Delphi survey results and attrition analysis is available from the author upon request.

**Appendix 13 Domain specific outcomes used for consensus surveys**

Due to space constraints, Appendix 11 containing the full list of outcomes used for consensus surveys is available from the author upon request.

**Appendix 14 Consensus survey rankings**

Due to space constraints, Appendix 14 containing the consensus survey rankings is available from the author upon request.

**Appendix 15 Consensus meeting 1 and 2 reports**

Due to space constraints, Appendix 15 containing the consensus survey reports is available from the author upon request.
